# The confounding effects of skin colour in photoacoustic imaging

**DOI:** 10.1101/2025.03.28.25324605

**Authors:** Thomas R. Else, Christine Loreno, Alice Groves, Benjamin T. Cox, Janek Gröhl, Inés Modolell, Sarah E. Bohndiek, Amit Roshan

## Abstract

Skin colour is known to confound readouts from optical devices that make measurements through the skin, which can adversely impact the care of patients with darker skin. Photoacoustic imaging (PAI) is making its way from the laboratory to the clinic, however, combining optics and ultrasound for deep tissue imaging leads to a complex relationship between photoacoustic-derived imaging biomarkers and skin melanin concentration. Furthermore, no generalisable correction of the confounding effects of skin colour in PAI has been demonstrated. We sought to overcome this limitation by recruiting a healthy volunteer cohort with the most diverse range of skin tones ever assembled in the field, with participants from Fitzpatrick types I to VI and with vitiligo. From this comprehensive dataset, we identified and characterised two physical mechanisms responsible for skin colour-dependent degradation in both image quality and biomarker quantification. Accompanied by detailed theoretical modelling, we demonstrated that strong light absorption by melanin leads to spectral colouring, which dominates in individuals with low skin melanin pigmentation. We further identified the backscattering of ultrasound waves generated in the skin as a major source of image artefacts for individuals with high skin melanin pigmentation. With this improved understanding of the physical basis, we were able to develop a fast and practicable correction method for spectral colouring and adapted a plane-wave ultrasound reconstruction algorithm to reveal the ultrasound scatterer distribution encoded in the photoacoustic timeseries. Our findings highlight the need for more advanced image reconstruction methods to enable equitable clinical application of PAI.

**One Sentence Summary:** Photoacoustic imaging is proven to suffer from measurement inaccuracies in people with darker skin, which could adversely impact patient care if not appropriately corrected.

## INTRODUCTION

The non-invasive measurement of blood oxygen saturation through the skin with optical technologies, such as pulse oximetry, is now widely acknowledged as being confounded by skin colour (*1, 2*). Melanin pigmentation in the skin strongly absorbs incident light, particularly at shorter wavelengths (*3*), which distorts the incident spectrum in a process known as ‘spectral colouring’. In pulse oximetry, given the spectral properties of melanin and oxy- and deoxy-haemoglobin, the dominant effect of light absorption by skin melanin is an overestimation of the blood oxygen saturation readout, particularly at dangerously low blood oxygen levels (*1*). Such overestimation in pulse oximetry can lead to missed hypoxaemia, which is recognised to have adversely affected patient management globally during the Covid-19 pandemic (*4*). Subsequently, several government reviews (e.g., in the United States (*5*) and the United Kingdom (*6*)) have highlighted the need to evaluate potential healthcare inequities in optical technologies that make measurements through the skin, and regulatory requirements have been updated to require that testing is conducted on participants with a diverse range of skin tones (*5*).

Photoacoustic imaging (PAI) unites light and sound to enable non-invasive deep tissue imaging. Commercially-available PAI systems typically exploit the lower scattering of near-infrared light to allow deep tissue imaging oxy- and deoxy-haemoglobin, lipids, and water (Fig. 1A). Performing PAI at multiple wavelengths enables the optical absorption of these different biomolecules to be resolved and processed into output biomarkers, such as haemoglobin concentration and blood oxygen saturation. PAI has been evaluated across numerous large-scale multi-centre clinical trials and has been shown to hold substantial promise for clinical management of a range of diseases, from inflammatory bowel diseases to breast cancer (*7*). Despite recent regulatory approvals in Europe, the USA, and Japan, a comprehensive assessment of the confounding effects of skin colour has yet to be performed for PAI.

**Fig. 1.**
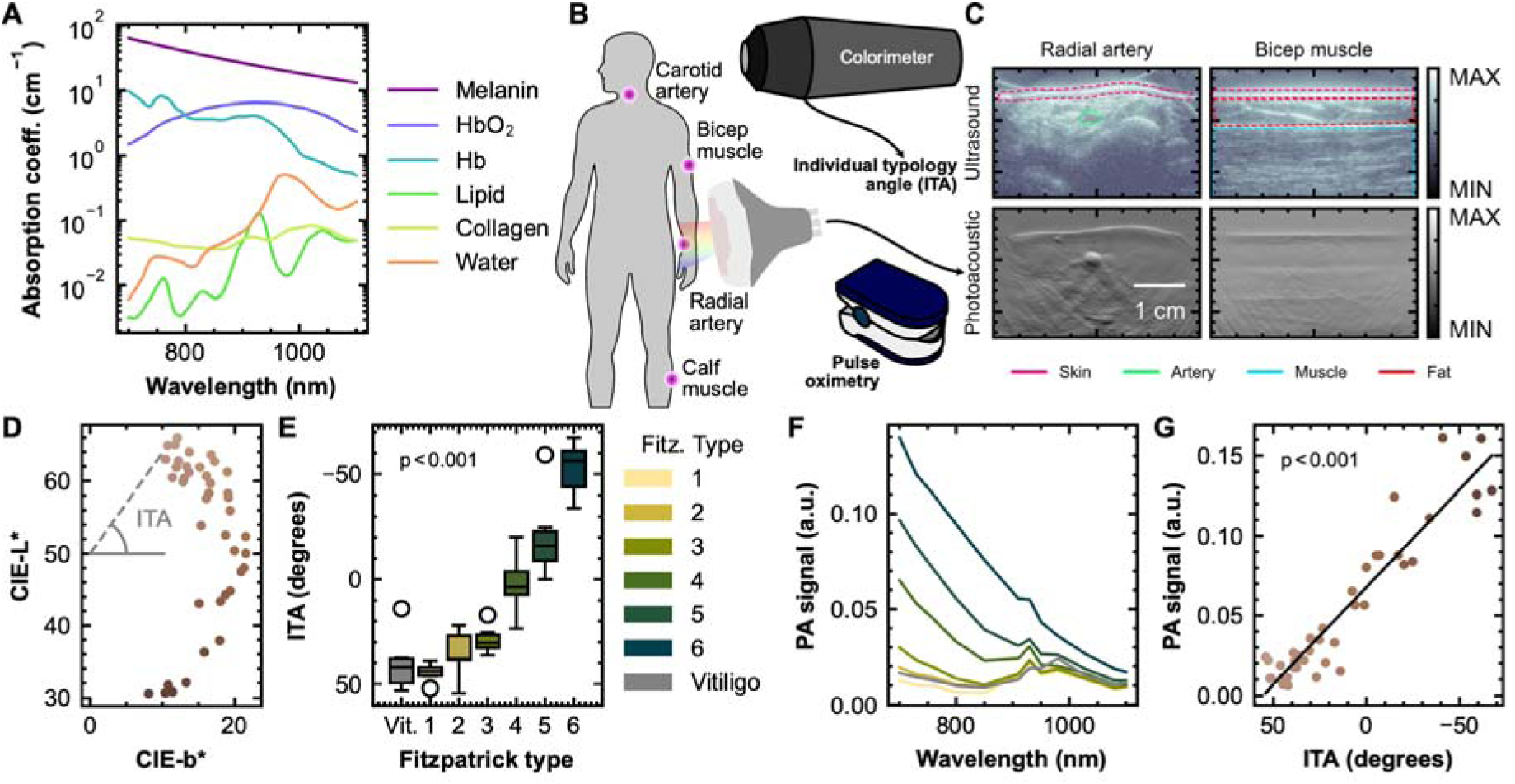
Photoacoustic signal amplitude correlates with subjective and quantitative measurements of skin colour, individual typology angle (ITA) and Fitzpatrick type. (**A**) Optical absorption spectra of dominant chromophores in the near infrared (*3*). Melanin spectrum shown at 40 % melanosome volume fraction (upper end of physiological levels), haemoglobin spectra are shown at the concentration found in blood, lipid collagen and water are shown in their pure forms. (**B**) Schematic overview of the study, highlighting imaging locations, colourimeter measurement of individual typology angle (ITA) and pulse oximetry. (**C**) Examples of photoacoustic and ultrasound images in the radial artery and bicep muscles highlighting key anatomical structures. (**D**) Plot of colourimeter measurements in CIE colour space, lightness L*, vs yellow-blue b*. (**E**) ITA from a representative region (neck) correlates with Fitzpatrick type. (**F**) The mean photoacoustic signal in a representative location (neck) from the skin surface across Fitzpatrick type groups. (**G**) 700 nm photoacoustic signal in the skin surface region (neck) correlates with ITA. Scatter plot markers in **D** and **G** are red/green/blue renderings of the colourimeter measurements.

The absorption coefficient of melanin in the epidermis can be higher than that of all other contributors for those with darker skin (Fig. 1A). Initial research efforts (*8–10*) suggest that skin colour can affect both image quality and biomarker quantification. Artefacts, known as clutter, increase in subjects with darker skin, obscuring the desired signal from features like blood vessels, whilst measurements made across multiple wavelengths demonstrate the familiar spectral colouring seen in other oximetry measurements, potentially introducing systematic biases in quantification of imaging biomarkers such as blood oxygen saturation (*8, 11, 12*). Increased clutter and spectral colouring could adversely affect image interpretation in the clinic, particularly in applications such as breast cancer where image feature sets are used to interpret the findings and provide disease staging information (*13–15*).

Accurate quantification in PAI is challenging even without the confounder of melanin (*16–20*). The challenge arises because the amount of light absorbed by a molecule, and hence the photoacoustic signal intensity, depends not only on the concentration of each absorbing molecule, but also on the spatially-varying light fluence (*21*). Inverting this process to obtain molecule concentrations and light scattering coefficients from photoacoustic images is impossible without making several assumptions (*22*). To avoid this complexity, an approximate linear spectral unmixing approach (*23*) is typically used in clinical applications (*24–26*), however, skin pigmentation would distort the calculation. To date, clinical studies have yet to report the impacts of skin tone on their linear spectral unmixing results, nor have they recruited diverse testing cohorts, while emerging preclinical studies of skin tone in PAI show inconsistent results (*9, 11*).

Here, we present the first comprehensive assessment of the confounding effects of skin tone in PAI. We conducted a volunteer study of participants with a diverse range of skin pigmentations and with vitiligo. We began by analysing the corruption of image quality and biomarker quantification with increasing skin melanin pigmentation. We then undertook detailed theoretical modelling to improve understanding of both the optical and acoustic effects observed. By modelling wavelength-dependent changes in light fluence as a function of skin pigmentation, we demonstrate a fast, practical and generalisable correction method for subjects with lower skin pigmentation. For individuals with very high skin pigmentation, image clutter is shown to originate from the skin acting as an ultrasound transducer, with photoacoustic waves generated in the skin being backscattered throughout the image field. We adapt a plane wave ultrasound reconstruction algorithm to invert the scattering problem, remarkably reconstructing images that are practically identical to conventional ultrasound images. We underscore the importance of PAI clinical trials testing on diverse populations and accelerating co-design of hardware and software to ensure equitable future application of PAI in the clinic.

## RESULTS

### A diverse healthy volunteer study was vital to understand confounding effects of skin colour in photoacoustic imaging

In our healthy volunteer study, we pre-screened potential participants using a questionnaire to establish their Fitzpatrick type, enabling fast and easy remote self-assessment of their qualitative skin tone grouping (*27*). To avoid other potential confounders, we sought to balance age, sex and body mass index between groups (summarised in Table 1, full data in Table S1). We recruited 6 participants from each Fitzpatrick type and 6 people with vitiligo for a total of 42 participants.

**Table 1.**
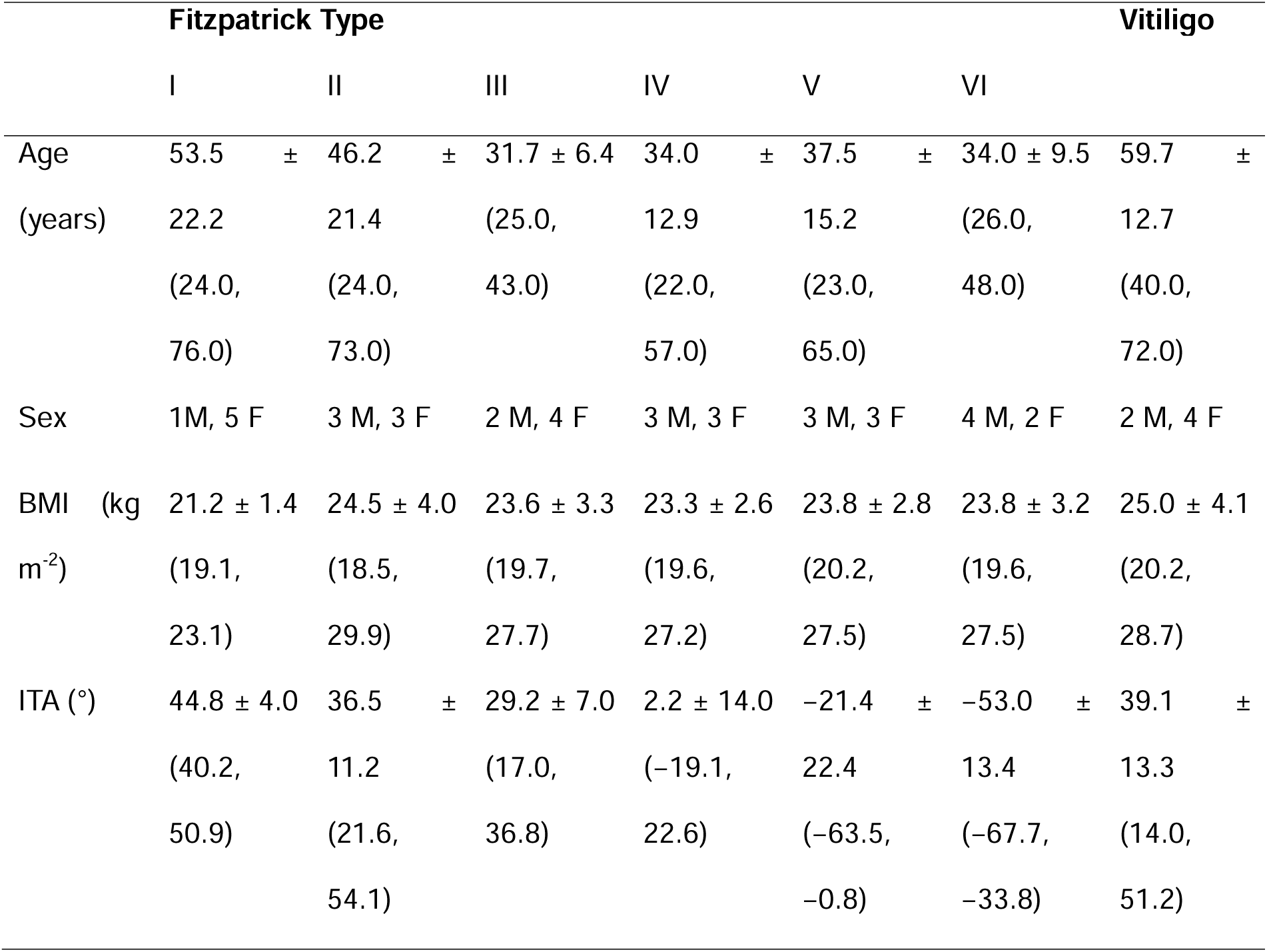
Summary of the study cohort. Numerical values are shown as mean ± standard deviation, with range in brackets. Total of six participants in each group (column).

All participants underwent photoacoustic imaging (PAI) with simultaneous reflection ultrasound (US) computed tomography and colourimetry at the imaging sites, as well as systemic pulse oximetry (Fig. 1B). The photoacoustic probe was positioned using the anatomical information from the ultrasound image and data acquisition was performed for 30 seconds per site (Fig. 1C). To maximise generalisability of findings to a broad range of clinical applications, we acquired PAI data in several anatomical sites, targeting major blood vessels and muscles, across 13 wavelengths, from 700 nm to 1110 nm (see Materials and Methods).

Colourimeter measurements were mapped to individual typology angle (ITA), a widely used measure of skin melanin level, which decreases with increasing melanin concentration (Fig. 1D). ITA decreased significantly with increasing Fitzpatrick type (p < 0.001 with a linear model), as observed in prior studies (*28, 29*), suggesting that the Fitzpatrick scale was a suitable proxy for skin colour to streamline participant recruitment (Fig. 1E).

We confirmed the relationship between photoacoustic signal from the skin surface and skin pigmentation quantified by colorimetry. Interestingly, we observed that the photoacoustic time series signal amplitude reached the upper limit of the analogue to digital converter in subjects with ITA as high as 40 (Fig. S1), resulting in clipping; band-pass filtering was applied prior to image reconstruction to reduce the effect. After model-based image reconstruction, the photoacoustic signal in the skin was obtained by drawing a polygonal region of interest (ROI) on the US images and applying it to the photoacoustic images (Fig. 1). Data from one participant was excluded, as ultrasound data was unavailable. The skin PA spectra showed the characteristic decrease with increasing wavelength associated with melanin absorption (Fig. 1F) and the PA signal at 700 nm correlated strongly with ITA (p < 0.001, linear model; Fig. 1F).

### Increasing skin pigmentation leads to extensive acoustic clutter artefacts

We first qualitatively assessed the appearance of the image features and artefacts across the dataset. In forearm scans, the radial artery was clearly distinguished at long wavelengths in all participants but was only visible at all wavelengths in participants with lighter skin tones (Fig. 2A). Conversely, it was entirely obscured by clutter (very dark or very light arc-shaped features below the skin surface in the images) in participants with darker skin, particularly at shorter wavelengths (Fig. 2A). Likewise, in scans of the bicep and calf muscles (Fig. S2 and S3), photoacoustic signals can distinguish the muscle from the subcutaneous fat in participants with lighter skin, but acoustic clutter comes to dominate in participants with darker skin.

**Fig. 2.**
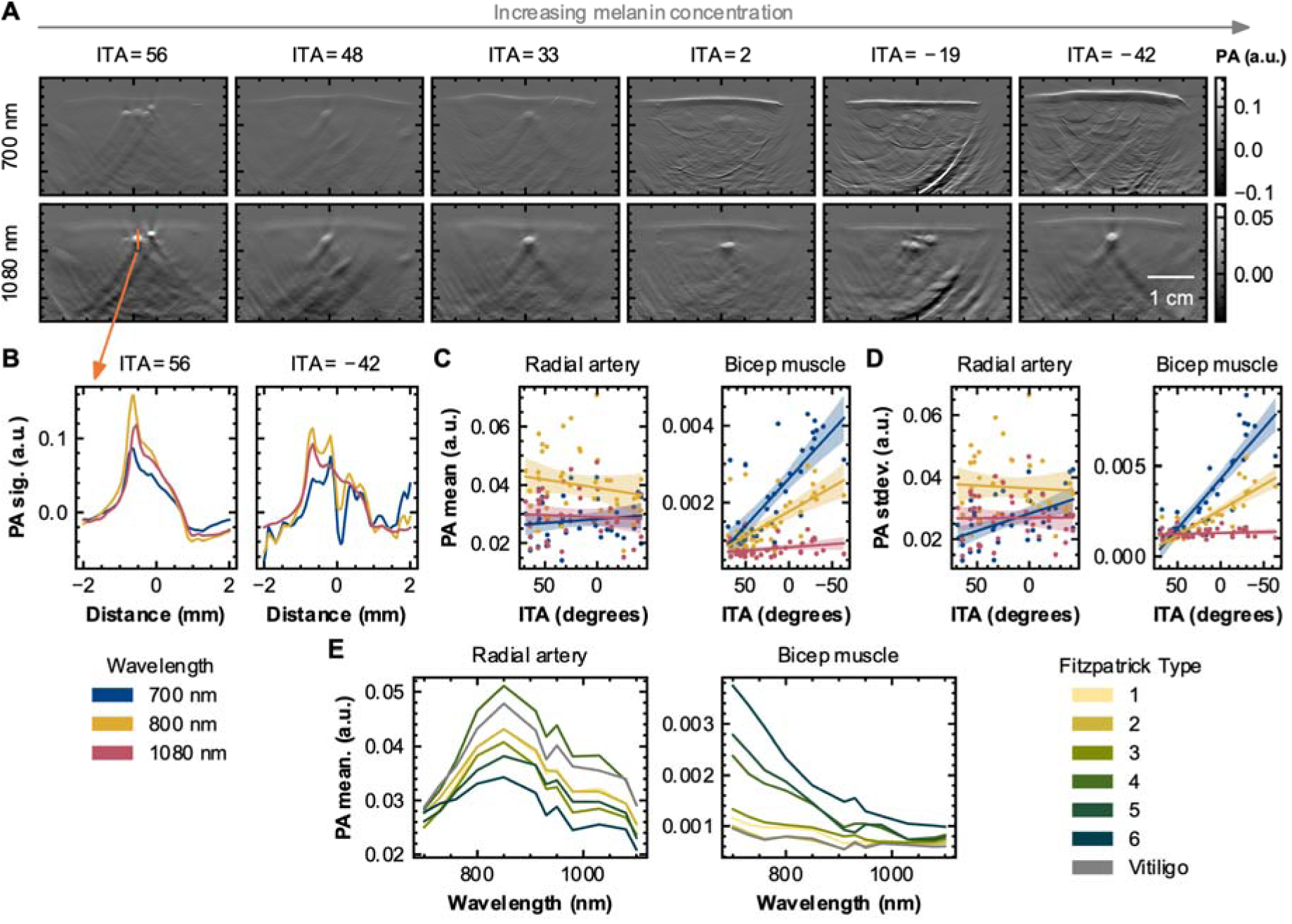
Single-wavelength photoacoustic data reveal increasing clutter signal with increasing skin pigmentation. (**A**) Example photoacoustic images of the radial artery at two wavelengths across a range of skin pigmentations, as measured with individual typology angle (ITA). (**B**) Vertical lines plots of PA signal intensity through the radial artery in the subjects with the maximum ITA (least melanin, left) and minimum (most melanin, right) at several wavelengths. (**C**) Mean photoacoustic signal in the radial artery and bicep at several wavelengths as a function of ITA. (**D**) The standard deviation of the photoacoustic signal, a measure of clutter, calculated across the radial artery and bicep muscle regions of interest as a function of ITA. Wavelengths shown in B-D are 700 nm (blue), 800 nm (orange) and 1080 nm (pink). (**E**) Mean photoacoustic spectrum for each Fitzpatrick type group and the vitiligo cohort in the radial artery and bicep muscle regions.

A bright horizontal artefact, located twice as deep as the muscle-fat interface, was observed in all muscle scans in subjects with darker skin, consistent with strong acoustic reflection of the photoacoustic wave at the muscle-fat interface (Fig. S2 and S3). Similar observations were made in scans of the neck, where veins around the carotid artery can be seen in photoacoustic images in almost all subjects, even at shorter wavelengths, but increased clutter is visible in subjects with darker skin, obscuring the carotid artery (Fig. S4).

Our qualitative assessment suggested that the forearm was the most informative site for understanding the confounding effects of skin colour due the presence of the radial artery as a focal feature, along with background tissue and bone. The remainder of the manuscript focuses primarily on analysis of the forearm, while the bicep is included as a comparative muscle.

We then performed a quantitative assessment of photoacoustic signals in the radial artery (forearm site) and muscle (bicep site). A vertical line plot through the radial artery illustrates the effects of acoustic clutter; in subjects with lighter skin, the radial artery has a consistent shape at all wavelengths, with sharply-defined edges, and a slight decay in intensity consistent with reducing light fluence (Fig. 2B). In subjects with darker skin, distinct oscillations could be observed in the line plots through the radial artery (Fig. 2B) due to stripe artefacts in the photoacoustic image and bands of low signal intensity. Perhaps surprisingly, the overall signal intensity of different structures did not decrease with increasing skin pigmentation, as might be naively predicted. Rather, in the radial artery, there was a negligible change in signal at several wavelengths used for blood vessel imaging in commercial devices (Fig. 2C, less than 1.3-fold changes, not statistically significant at 700 nm, 800 nm and 1080 nm from ITA = 68 to ITA = -42). The standard deviation of the signal across the radial artery increased significantly at 700 nm, in line with the observations of increased clutter (Fig. 2D, p < 0.001 at 700 nm, p = 0.70 at 800 nm, p = 0.95 at 1080 nm). The averaged photoacoustic spectrum of the radial artery had substantial variability across Fitzpatrick types, maintaining the broad shape associated with oxygenated blood, with a slight increase at low wavelengths at the highest Fitzpatrick types (Fig. 2E).

In the bicep, there was a dramatic increase in mean photoacoustic signal with increasing skin pigmentation, consistent with the line-shaped clutter observed in the images (4.8-fold from ITA = 70 to ITA = -63 and p < 0.001 at 700 nm, 3-fold and p < 0.001 at 800 nm, 1.3-fold and p = 0.08 at 1080 nm). A significant increase in signal standard deviation was observed with increasing skin pigmentation in the bicep muscle (p < 0.001 at 700 nm, p < 0.001 at 800 nm, p = 0.29 at 1080 nm, Fig. 2C). The overall photoacoustic spectrum across the muscle varied dramatically across Fitzpatrick types, moving from a relatively flat spectrum to one with substantial wavelength dependence, resembling the optical absorption spectrum of melanin (Fig. 2C).

### Estimates of blood oxygen saturation are confounded by skin pigmentation

We next applied linear spectral unmixing for oxy-(HbO_2_) and deoxy-haemoglobin (Hb) to our dataset at wavelengths between 700 nm and 900 nm and estimated both total haemoglobin (THb = HbO_2_ + Hb) and blood oxygen saturation (sO_2_^EST^ = HbO_2_ / THb) as commonly evaluated PAI biomarkers in the clinic. Encouragingly, HbO_2_ images showed blood vessels quite clearly, even in subjects with darker skin (Fig. 3A upper row). On the other hand, Hb images showed a high level of clutter with increasing melanin and poor blood vessel visibility (Fig. 3A lower row).

**Fig. 3.**
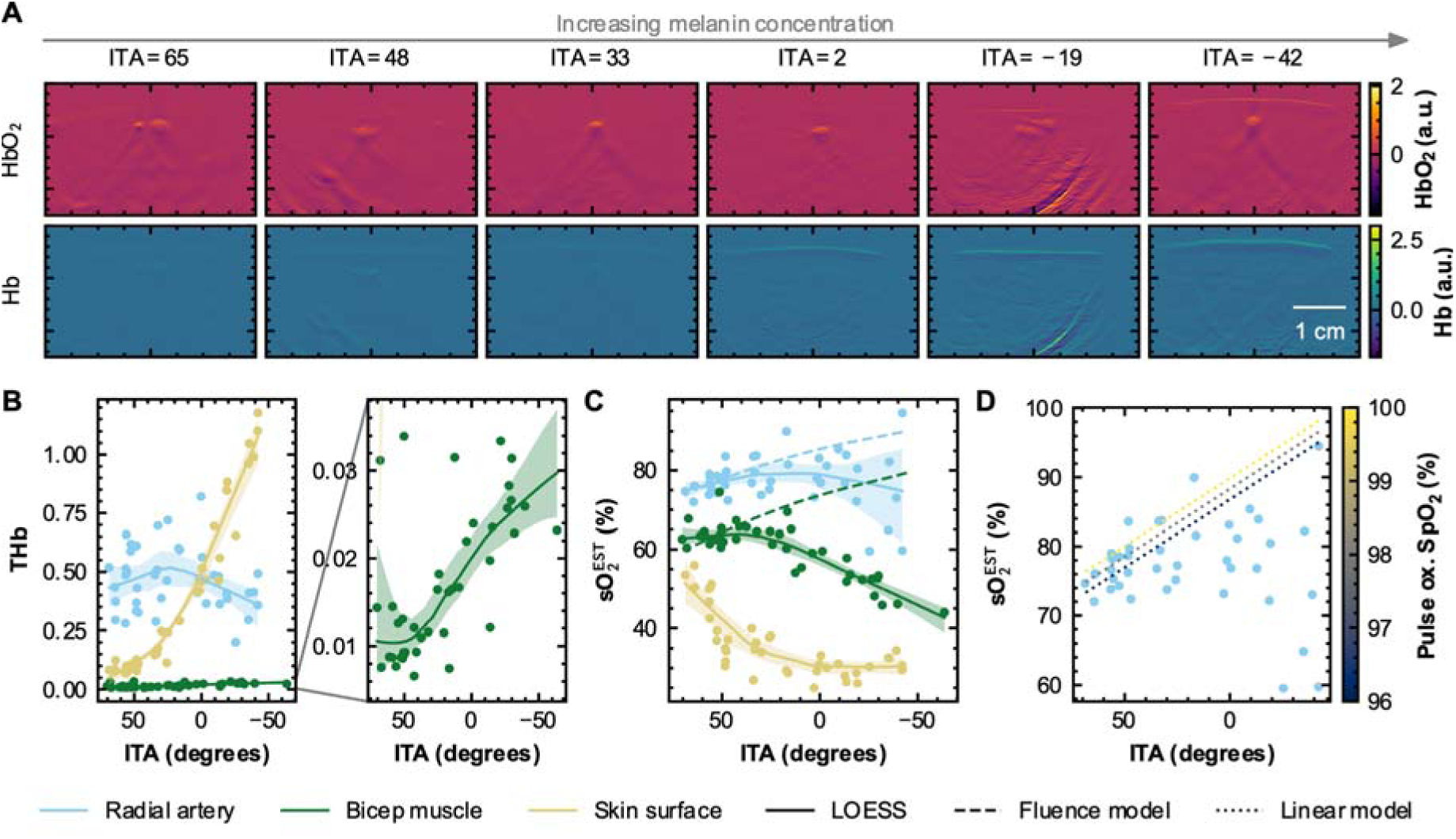
Linear spectral unmixing results in regions of interest show a strong dependence on skin pigmentation. (**A**) Unmixed oxyhaemoglobin (HbO_2_) and deoxyhaemoglobin (Hb) images of six participants across the range of skin pigmentations. Unmixed total haemoglobin (**B**) and blood oxygenation (sO_2_^EST^; **C**) variation in the radial artery, bicep muscle and skin surface regions of interest. LOWESS fits and 95 % confidence intervals are shown as solid lines and shaded regions respectively. A light fluence model relative to the minimum melanin level is shown as a dotted line (**B** and **C**). (**D**) A linear mixed effects model fit over the linear skin colour range (ITA > 10) shows the unmixed sO ^EST^ correlation with pulse oximetry. Dotted lines show the linear mixed effects model at 96, 98 and 100 % SpO_2_.

By extracting average values across ROIs, we were able to identify several important trends that indicate challenges in the clinical translation of PAI biomarkers. Estimates of total haemoglobin in the skin surface ROI increased dramatically as ITA decreased, highlighting a limitation of linear unmixing -- melanin absorption can be mistaken for deoxyhaemoglobin -- leading also to a decrease in sO_2_^EST^ with decreasing ITA (Fig. 3B, C). In the radial artery, there was a negligible change in THb with ITA (p = 0.30), but substantial variability between subjects (Fig. 3B). At low melanin levels (high ITA values), sO_2_^EST^ increased with decreasing ITA (p = 0.006, n = 25 subjects ITA > 10), while at high melanin levels (lower ITA values), the trend reverses and there is substantial variability between subjects (p = 0.39, n = 16 subjects ITA < 10) (Fig. 3C). In subjects with lower melanin levels (Fig. 3C, ITA > 10), sO_2_^EST^ correlated with pulse oximetry measurements (Fig. 3D, p = 0.051, coefficient = 0.73, [95 % CI -0.004, 1.463], linear mixed effects model, 163 observations from 18 participants). In the bicep muscle, there was a dramatic increase in THb with increasing melanin level (p < 0.001, linear model), consistent with the increase in clutter signal observed in single wavelength data (Fig. 3B zoomed graph). Again, sO_2_ ^EST^ follows a more complex trend, staying approximately level with increasing melanin at low melanin levels (ITA < 10, p = 0.57), before decreasing significantly at high melanin levels (ITA < 10, p < 0.001, [95 % CI 0.132, 0.306]; Fig. 3C).

To establish the basis for the dependence of sO_2_ ^EST^ on skin colour, we simulated the effect of spectral colouring from melanin on light fluence and the consequent effect on the resulting PAI sO_2_^EST^ (see Materials and Methods). The model had one free parameter, the sO_2_^EST^ at zero melanin concentration, which was dependent on the non-melanin absorption and scattering of the surrounding tissue. The theoretical model was found to fit very well at low melanin levels (ITA ∼> 30) in the bicep muscle and the radial artery (shown as a dashed line on Fig. 3C). Our fluence model fits particularly well for the radial artery, with a likelihood ratio of 0.016 compared to a constant model (ITA > 30). For the bicep muscle, the model is not statistically any better than a constant (likelihood ratio of 2.1) but is not inconsistent either (2-tailed t-test on residuals p-value = 0.89; ITA > 30). The model gradually diverges from the experimental data as melanin concentration increases, suggesting that spectral colouring ceases to be the dominant process at high melanin levels (Fig. 3C).

### Reproducibility was partially influenced by participant skin tone

To assess the potential impact of skin pigmentation on reproducibility of photoacoustic measurements, we evaluated the coefficient of variation of single-wavelength measurements and unmixed coefficients sO_2_ ^EST^ and THb across three scans, with replacement, in all participants. For comparative reference, a vendor-provided phantom was scanned prior to scanning each volunteer. The mean signal inside each phantom inclusion was evaluated over time, and the coefficient of variation was calculated (Fig. S5). In the phantom inclusion with the highest photoacoustic signal intensity, which is of the same order of magnitude as measurements in the radial artery, the coefficient of variation was below 10 % for all wavelengths below 1000 nm, and no higher than 11.5 % for any wavelength. The mean coefficient of variation across all wavelengths from 680 nm to 1100 nm, inclusive, was 6.8 %.

In measurements of the radial artery, there was a very slight decrease in the coefficient of variation with increasing melanin level (p = 0.019 and r = 0.41 at 800 nm, p = 0.049 and r = 0.35 at 1080 nm, Spearman’s rank (SR) test, Fig. S6A,B upper row). In the bicep muscle, there was no significant correlation between ITA and coefficient of variation (Fig. S6A,B lower row). Overall, across all wavelengths, there was a mean coefficient of variation of 22 % in the half of the cohort with lighter skin and 14 % in the half with darker skin. For THb, a similar observation was made, with a slight correlation between coefficient of variation and ITA in the radial artery (p = 0.0201, r = 0.39, SR test) and no correlation in the bicep muscle (p = 0.50, r = 0.12, SR test) (Fig. S7). Coefficient of variation in sO_2_ ^EST^ correlated significantly with ITA in the radial artery (p = 0.029, r = -0.37), but there was no trend in the bicep muscle data (p = 0.56, r = -0.10, SR test; Fig. S7). In the radial artery, the mean coefficient of variation for THb and sO_2_ ^EST^ was 25 % and 3.3 % respectively for the lighter half of the cohort and 17 % and 5.5 % for the half with darker skin. In the bicep muscle, the mean coefficient of variation for THb and sO_2_ ^EST^ was 24 % and 4.2 % respectively for the lighter half of the cohort and 27 % and 5.3 % for the half with darker skin.

### Photoacoustic imaging is sensitive to subtle changes in skin pigmentation in participants with vitiligo

Photoacoustic images from participants with vitiligo were compared between pigmented and non-pigmented (vitiligo affected) regions, as determined by white light and UV photography (Fig. 4A). All participants in the vitiligo cohort would otherwise have been placed in Fitzpatrick types I, II, or III, so there is relatively low variation in skin pigmentation in the cohort. Consequently, while increases in melanin absorption of the epidermis between pigmented regions and non-pigmented can be seen in 700 nm photoacoustic images, no changes in the influence of image artefacts can be observed (Fig. 4B). By comparison, ITA values had a highly significant difference between pigmented and non-pigmented patches (p < 0.001, linear mixed effects model; Fig. 4C). Photoacoustic signals from the skin surface region at the lowest wavelength of 700 nm (highest melanin absorption) increased only modestly between non-pigmented regions and pigmented regions, likely because of absorption from other chromophores like haemoglobin (p = 0.059, linear mixed effects model; Fig. 4D). There was a high correlation, however, between ITA and 700 nm skin surface photoacoustic signal, as observed in the wider cohort (p = 0.015, linear mixed effects model; Fig. 4D).

**Fig. 4.**
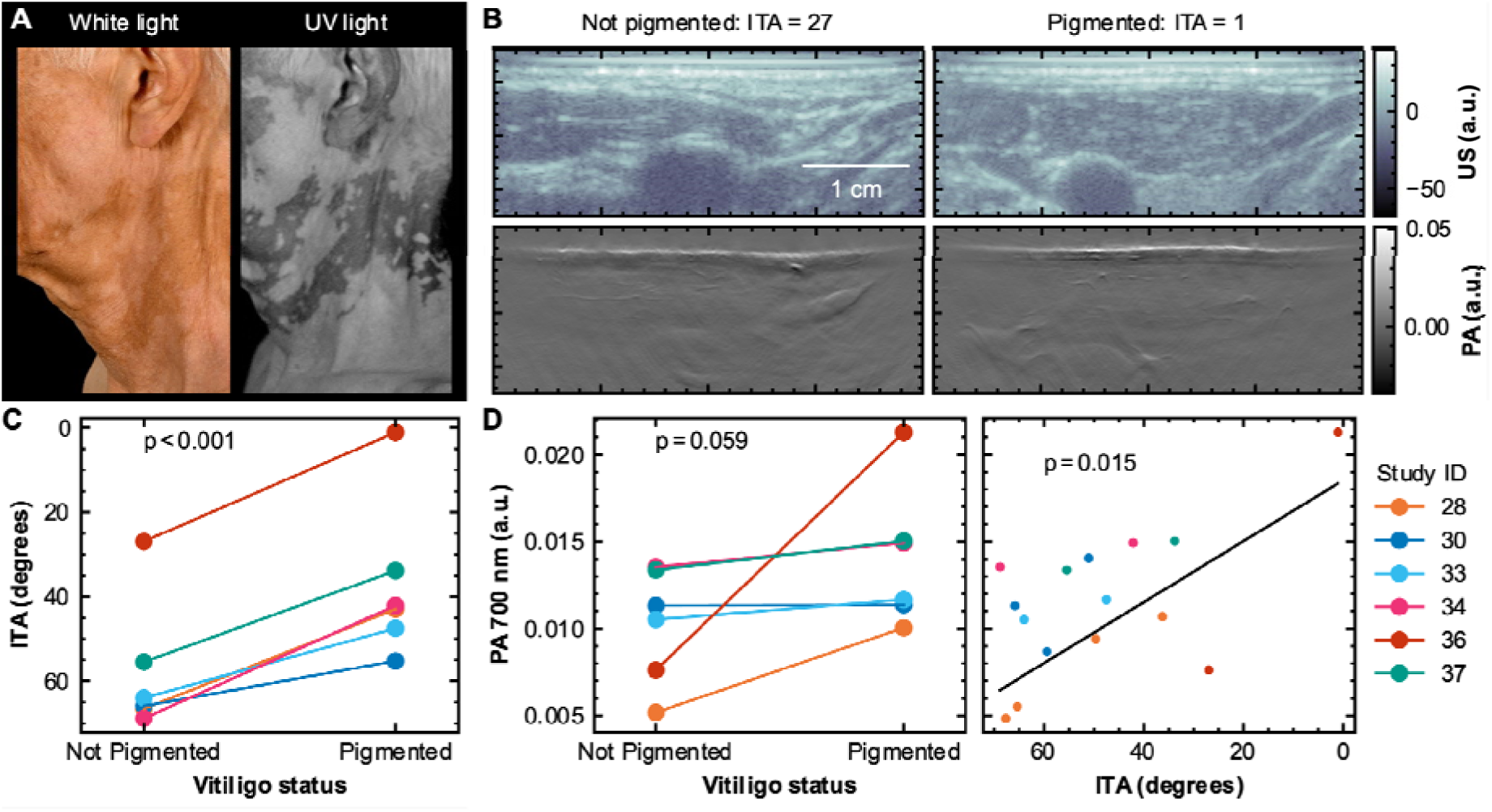
Photoacoustic imaging in subjects with vitiligo reveals high sensitivity to melanin concentration in the epidermis. (**A**) Example white light and UV light photographs of the neck of a participant with vitiligo. (**B**) Ultrasound (US) and 700 nm photoacoustic images of the neck (carotid artery) over a non-pigmented patch and a pigmented patch of the same participant. (**C**) Individual typology angle for each vitiligo participant over vitiligo patches (not pigmented) and patches with normal pigmentation (pigmented). (**D**) Mean 700 nm photoacoustic signal in the epidermis in vitiligo patches (not pigmented) and patches with normal skin pigmentation (pigmented). Plot of the same 700 nm photoacoustic signal against ITA with a linear mixed effects model line of best fit (black). All p-values shown were calculated using a linear mixed effects model.

### A fast generalisable method can correct for melanin spectral colouring and improve estimation of blood oxygen saturation in participants with low skin pigmentation

To mitigate the spectral colouring component of skin colour bias, we developed a fluence correction approach. We modelled light transport through a simple tissue model consisting of an epidermis with varying melanosome volume fraction, a blood vessel and background scattering and absorbing tissue (Fig. 5A; see Materials and Methods). The relationship between melanosome volume fraction and ITA was established in a previous study, enabling us to connect the simulated data to our *in vivo* measurements (*11*). The simulated illumination geometry was consistent with the clinical PA system used in this study. The simulated wavelength was varied, and the light fluence in the blood vessel was averaged to give a lookup table. The lookup table was normalised by the lowest melanin concentration simulated, and interpolated across wavelength and ITA, giving a correction factor that could be applied to the clinical data (Fig. 5B). The correction factor did not vary spatially, so was calculated by a simple cubic spline interpolation of the lookup table (Fig. 5C)

**Fig. 5.**
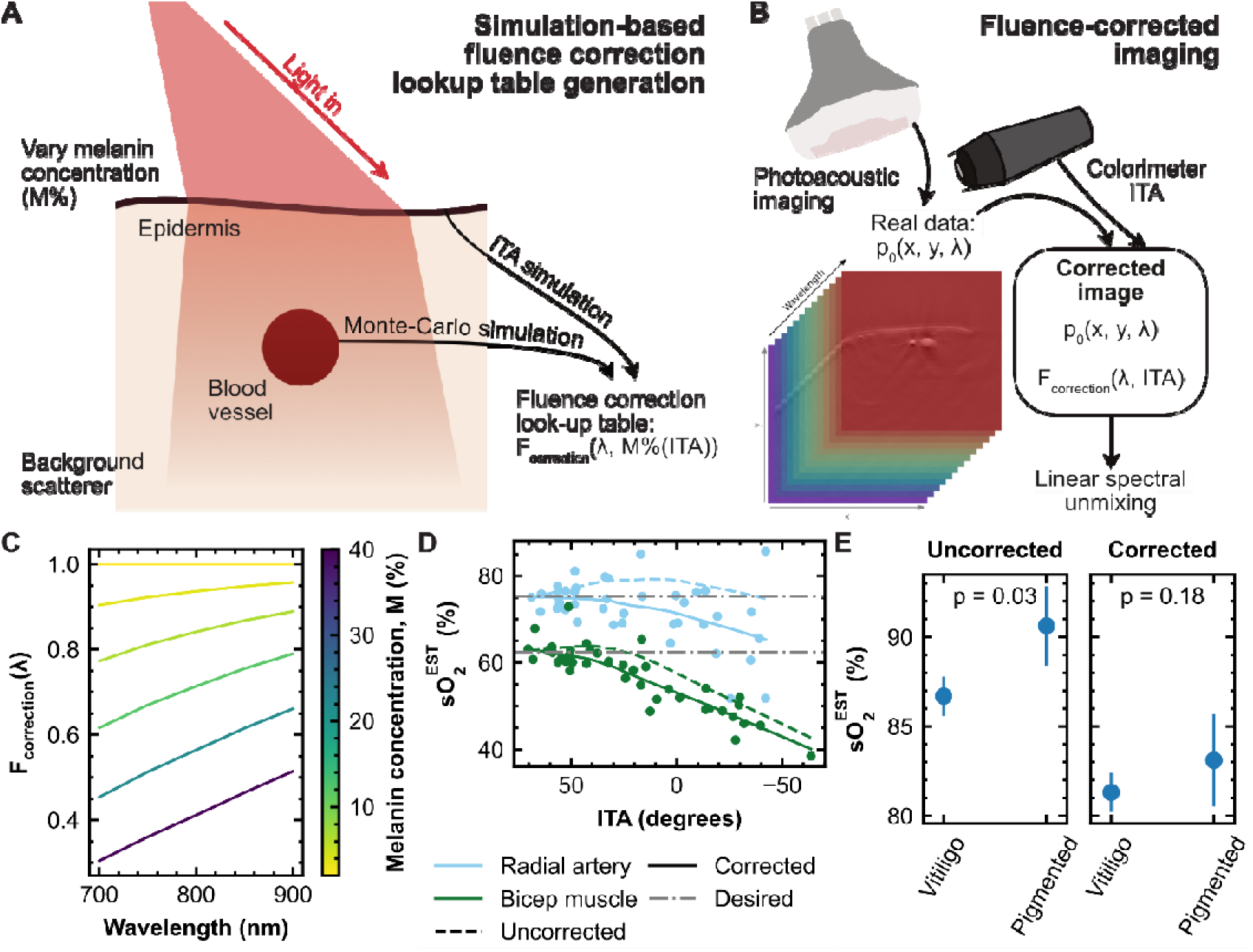
A Monte-Carlo simulation model for spectral colouring correction due to melanin in the epidermis. (**A**) A schematic of the approach taken to fluence correction, where melanosome volume fraction (M %) is related to light fluence (F) and individual typology angle (ITA) through optical modelling. (**B**) A schematic of how the fluence correction is applied to the real photoacoustic data. (**C**) The normalised fluence correction factor as a function of wavelength and melanosome volume fraction (M %). (**D**) Blood oxygenation estimates from linear unmixing in the radial artery and the bicep muscle of healthy volunteers before and after fluence correction, with a LOWESS trend line. (**E**) Blood oxygenation estimates from linear unmixing of the carotid artery from a single participant with vitiligo over a non-pigmented (vitiligo) patch and a pigmented patch, before and after correction; p-values show a 1-tailed t-test.

The correction approach was then tested on the reconstructed photoacoustic images acquired in healthy volunteers, which were divided by the associated correction factor to give a corrected image. Linear spectral unmixing was then applied to estimate blood oxygenation (Fig. 5D). The increase in sO_2_^EST^ with increasing skin pigmentation associated with spectral colouring was normalised in the corrected images at low melanin levels. The correction method also worked well for a matched region on a vitiligo participant and was able to match the sO_2_^EST^ across their vitiligo and pigmented regions. The correction factor removed the statistically significant relationship between ITA and sO_2_^EST^ of the radial artery in subjects with ITA > 10 (p = 0.72).

### Acoustic clutter arises from the skin, which acts as a secondary source of ultrasound in participants with high skin pigmentation

Having established how changes in the skin pigmentation affect the propagation of light in the tissue and subsequently corrupt estimation of THb and sO_2_^EST^, we finally sought to develop a deeper understanding of the origin of the clutter artefacts observed. Initial qualitative evaluation suggested that the artefacts appeared in the PA image approximately twice as far below the skin surface as strongly reflecting structures in the ultrasound image, with spectral features similar to that of melanin in the skin surface (Fig. 1E, Figs S2 and S3). The consistent appearance suggested that the skin could be acting as a secondary source of acoustic waves due to the strong optical absorption, which were backscattered at interfaces between tissues of different acoustic impedance. Upon further interrogation, it was found that the backscattered signals were being erroneously reconstructed into the photoacoustic image based on the wave travel time (Fig. 6A). Such artefacts could easily be mistaken as low-oxygenation blood vessels, which could be highly misleading in a clinical assessment.

**Fig. 6.**
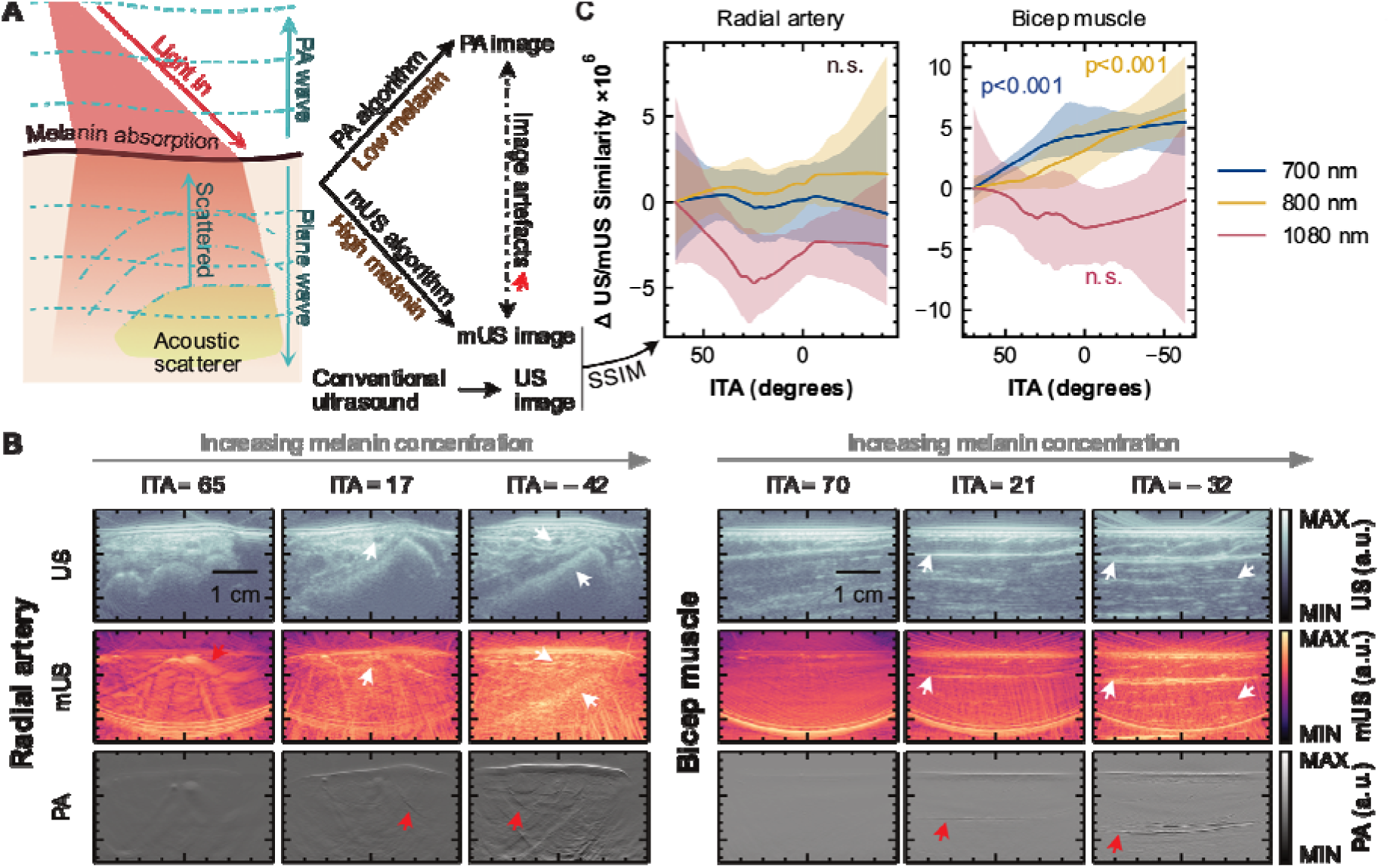
Plane wave ultrasound reconstruction illustrates that ultrasound scattering artefacts dominate in participants with darker skin pigmentation at shorter wavelengths. (**A**) A schematic showing the generation of ultrasound plane waves in the skin surface, which scatter off deeper scattering tissues, like bones and muscles. A standard photoacoustic (PA) algorithm generates a good PA image, whilst the melanin ultrasound (mUS) algorithm generates an ultrasound (US) image at high melanin levels. At the other extremes, mUS and PA images contain artefacts (shown with a red arrow). (**B**) Photoacoustic (PA), melanin ultrasound (mUS) and conventional ultrasound (US) images at low, medium and high melanin levels from the radial artery (left) and bicep muscle (right). Features visible in both mUS and US images are highlighted with a white arrow, while artefacts are highlighted by a red arrow. (**C**) A plot of a LOWESS fit of structural similarity index metric (SSIM) change between mUS and conventional US images as a function of ITA in all subjects at several wavelengths, relative to the value at maximum ITA; shaded areas show 95 % confidence intervals for the fit. Quoted p-values use the Spearman’s rank test.

To further illustrate the source of acoustic clutter, we developed an algorithm that was able, in the high melanin limit, to recover the spatial ultrasound scattering distribution from the clutter. The resulting images are referred to as melanin ultrasound (mUS). We adapted an ultrasound reconstruction algorithm originally used for plane wave optical ultrasound (*30*), making changes to account for the arc-shaped transducer and the spatial offset between the detector and the epidermis source of ultrasound (see Materials and Methods). We validated our algorithm with computational mUS and US simulations, showing that mUS images resemble conventional US in the high melanin limit (Fig. S8).

Remarkably, mUS images computed from 700 nm PA time series data resemble conventional ultrasound images in all imaging sites, even at only modest levels of melanin (Fig. 6B). At very high melanin levels, deep anatomical structures, such as the radius bone, striations in the muscles, and the carotid artery can be visualised in mUS images (Fig. 6B, Figs S9 and S10). By contrast, mUS images at low melanin levels contain artefacts from features like blood vessels and muscles, as the ultrasound wave generated in structures like the radial artery and bicep muscle outweighs the secondary melanin absorption wave (Fig. 6B, S9 and S10). The structural similarity index metric (SSIM) was calculated to compare mUS and US images (Fig. 6C). At shorter wavelengths, SSIM shows a significant correlation with ITA in the bicep muscle (p < 0.001 for 700 nm and 800 nm, p = 0.48 for 1080 nm, Spearman’s rank test), but not in the radial artery (p > 0.5 at 700 nm, 800 nm and 1080 nm, Spearman’s rank test). Our observations suggest that the epidermis acts as a strong source of ultrasound waves, overwhelming primary PA signals at high melanin concentrations, imprinting artefacts with a melanin spectral fingerprint on PA images. Thus, our previous observations of sO_2_^EST^ diverging from the fluence model at high melanin levels can be attributed to acoustic heterogeneity, a factor not included in the PAI reconstruction model, which assumes a constant speed of sound.

## DISCUSSION

PAI holds potential for adoption in clinical indications ranging from peripheral artery disease assessment to cancer diagnosis, but the confounding effects of skin colour on PAI quality and biomarker measurements have not yet been well established. We carried out a healthy volunteer study in 42 participants across a wide range of skin tones, including 6 participants with vitiligo. We showed that single wavelength measurements and the linear unmixing quantities THb and sO_2_^EST^ are all affected by skin pigmentation in a non-linear manner due to confounding effects arising from both optical (spectral colouring) and acoustic (backscattering) effects. We revealed the physical mechanisms behind the observed effects and took steps towards potential correction methodologies through comprehensive computational modelling.

To address the spectral colouring effects, we related a computational model of light transport to an independent measure of skin colour, ITA, and were successfully able to predict the changing light fluence spectrum below the epidermis, giving improved quantitative oximetry values at low melanin levels. To address the acoustic effects, we modelled the epidermis as a source of ultrasound plane waves, allowing us to reconstruct ultrasound images from the clutter signal. Our work paves the way for skin colour independent clinical photoacoustic imaging, addressing some challenges, but revealing the difficulties of tackling this issue in a universal manner.

Our observations suggest that optical and acoustic phenomena separately contribute to photoacoustic skin tone dependence. Optical effects from melanin absorption change the light fluence in deeper tissue regions, like muscles or blood vessels, which give a misleading spectral appearance. At lower melanin levels (ITA ∼>10), our observations are consistent with previous theoretical work, which predicted an increase in unmixed blood oxygenation with increasing melanin (*11*). Yet the optical model breaks down at higher melanin levels. Above ITA ∼ 10, we saw an unexpected rise in single wavelength photoacoustic signal and a decrease in unmixed blood oxygenation. We were able to attribute the divergence from our optical model to backscattering of photoacoustic ultrasound generated in the epidermis, and the constant speed of sound assumption in image reconstruction. The resulting artefacts have the same spectrum as melanin and obscure the desired primary photoacoustic signal.

Having observed these confounders, we then sought to correct them. We showed that a light fluence model can partially compensate for skin colour corruption of sO_2_^EST^. The correction method did not depend on any empirical parameters; we directly modelled the illumination geometry and individual typology angle as a function of melanosome volume fraction. We chose not to derive an empirical correction factor for sO_2_^EST^ correction, instead focussing on the optical problem, as differences in tissue acoustic properties would alter the clutter level, leading to more or less spectral corruption in different anatomical sites. Our proposed correction model, therefore generalises to other tissue types, like muscles, while previous empirical formulae were derived specifically for arteries (*9*). The model also takes ITA as input, rather than a qualitative factor, like Fitzpatrick type, allowing for skin colour differences within the same subject to be modelled, neatly demonstrated by data in a subject with vitiligo. The lookup-table approach would easily integrate with existing clinical software without demanding computational overheads and could perhaps even derive ITA directly from the photoacoustic image given how well correlated we demonstrated them to be, eliminating the need for an independent colourimeter.

The acoustic effect means that photoacoustic images of participants with higher melanin pigmentation (ITA ∼< 10) are not, in fact, what we usually think of as photoacoustic images. Rather, the time series data is far more like a plane wave ultrasound scan. Because of the high optical absorption of melanin, the skin surface acts as an ultrasound transmitter, emitting waves into the deeper tissue, which give rise to reflected ultrasound signals that are far stronger than the desired primary photoacoustic signal from the tissue of interest. Remarkably, we were able to generate excellent ultrasound images from the PA time series data, convincingly illustrating how dominant backscattering is. Unfortunately, this mapping approach is not a correction for the confounder. Rather, it demonstrates the severity of the problem and indicates ways in which it might be addressed in future work. For example, knowing that the ultrasound reflectivity of the tissue can be obtained from photoacoustic time series data suggests that it could be incorporated into a model to separate out the primary photoacoustic signal from the scattered signals. Similar challenges are observed in other areas of photoacoustic imaging, such as in the brain, where jointly reconstructing initial pressure and speed of sound have shown promise (*31*). Furthermore, since the scattered signal has the same wavelength dependence as the skin surface, a spectral unmixing-like approach might enable some bias reduction.

This is the largest healthy volunteer study of its kind so far in PAI, giving several key new insights into the phenomenon of skin colour bias in photoacoustic imaging. There remain limitations, however, that could be addressed in future work. The study was carried out only with a single photoacoustic imaging system, with an arc-shaped detector, 2D acquisition geometry and a relatively wide detection bandwidth. How these results would translate to different systems, particularly those with different illumination and acoustic detection hardware remains unclear. In particular, the clutter signal will depend on transducer characteristics, such as bandwidth and noise level. Likewise, the lookup table generated for melanin absorption fluence correction will depend on the illumination geometry and must be recalculated and validated on other photoacoustic systems before it can be used in clinical practice. Furthermore, this study did not modulate the photoacoustic signal, as sometimes done in clinical photoacoustic applications, for example, with a blood pressure cuff or exercise. One would expect that “delta” metrics would be less susceptible to skin colour bias, as artefacts would remain relatively constant over the perturbation.

The competing effects of the acoustic and optical confounders could have profound implications for clinical translation of photoacoustic imaging. Either confounder may dominate depending on the subject’s skin colour, making blood vessels appear more or less oxygenated than they truly are depending on the subject’s precise skin colour. Perhaps more concerningly, artefacts may resemble hypoxic blood vessels, whose intensity depends not primarily on optical properties as we tend to expect, but on acoustic properties like density and elasticity of the surrounding tissue. Such confounders, if not accounted for, could have alarming consequences in applications like breast cancer diagnosis, for which regulatory approval has already been achieved, and where blood oxygenation is a key marker of cancer stage (*14, 15*). It is incumbent on the photoacoustic community to tackle this problem before widespread adoption of the technology.

## MATERIALS AND METHODS

To examine the effects of skin colour on PAI, a prospective, exploratory observational trial was conducted (PAISKINTONE). PAISKINTONE was approved by the East of England - Cambridge South Research Committee (Ref: 23/EE/0019) and conducted in accordance with the Declaration of Helsinki. Written informed consent was obtained from all study participants. The trial was registered with clinicaltrials.gov: https://clinicaltrials.gov/study/NCT05554523. The study started in June 2023 and finished recruitment in August 2024. A total of 42 participants were recruited for the study. The participants had a gender distribution of 24 females and 18 males (Table 1). Participants were recruited by flyers within the University of Cambridge, through invitation by the dermatology clinic of Cambridge University Hospital NHS Foundation Trust, or through invitation by the Alliance for Cancer Early Detection (ACED) cohort.

Participants were eligible if they were between the ages of 20 and 80, could provide informed written consent in English, and had a body mass index (BMI) of between 18.5 and 29 (not obese or underweight). Participants were excluded if they had tattoos or scarring over the imaging regions or if they had pulmonary sleep disorder or any other active respiratory condition that could affect blood oxygenation. Throughout the procedure, participants were monitored for adverse events, such as unacceptable skin erythema, at which point they would have been withdrawn. No adverse events had been observed as of the time of writing. Participants were free to withdraw at any point.

Prior to the study visit, potential participants were screened to assess their eligibility and Fitzpatrick skin type assignment. The Fitzpatrick skin type was identified by a questionnaire to ensure that we recruited a maximum of 6 participants for each group. While the Fitzpatrick skin type was originally designed as a method for categorising a person’s risk of skin cancer (*32*), it has since been used in several studies as a proxy measure of skin colour. It does not consider differences in skin colour between different positions of the body or changes over time (e.g. because of vitiligo or sun exposure) therefore it was used here solely as a straightforward method for ensuring that a representative, diverse cohort of participants were recruited.

Participants were invited for a single visit of approximately 1 hour duration. At the start of the visit, written informed consent was obtained, height and weight were measured, and eligibility was confirmed. The desired imaging sites were shaved if necessary to ensure good acoustic coupling between the photoacoustic probe and the skin.

Colourimeter measurements (Colorimeter DSM-4, Cortex Technology, Aalborg, Denmark) were taken at each imaging site, giving a quantitative measure of skin colour. The colour measurement is based on diffuse reflectance spectroscopy in a 45°/0° configuration. The colourimeter returns the reflected colour in several colour space representations (CIE-L*a*b*, CIE-L*C*h, CIE-XYZ) and several derived quantities, including the individual typology angle (ITA, Equation 1), which has been proposed as a quantitative measure of skin colour. All specular reflections were excluded from the measurement. Measurements were performed within an 8 mm diameter opening in the front of the device. The colourimeter was calibrated daily, using dark, white and gloss references provided by the manufacturer.

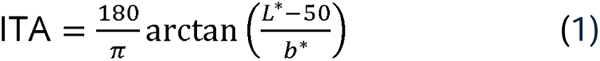

Pulse oximetry blood oxygen (B125 Patient Monitor, GE Health, Wisconsin, United States) saturation readings were recorded at the start of each scan. The pulse oximeter was used to give an approximation of the oxygen saturation of the arterial blood in the finger.

### Photoacoustic imaging

PAI was conducted using the MSOT Acuity Echo CE system (iThera Medical GmbH, Munich, Germany). Procedures were conducted under laser safety approved conditions. Light excitation pulses were provided by a tuneable optical parametric oscillator (OPO) pumped by a neodymium-doped yttrium aluminium garnet (Nd-YAG) laser, with a pulse repetition rate of 25 Hz. The laser output wavelength can be tuned from 660 nm to 1300 nm. Laser light was delivered to the probe by a single optical fibre. The output energy of the laser was less than 30 mJ, falling below the maximum permissible exposure limits. The laser energy of each pulse was measured automatically by the system.

The photoacoustic signals were detected by a two-dimensional array of ultrasound transducers arranged in an arc shape within the probe (central detection frequency 4 MHz, 256 detector elements). The resulting images have a field of view of 30 mm and spatial resolution 150 µm. Reflection ultrasound computed tomography images are provided by the system in parallel, giving intrinsically co-registered anatomical information alongside the functional information provided by PAI. The ultrasound images were used to position the probe, as they provided much clearer anatomical information than photoacoustic images.

PAI was carried out on several sites in the body. Clear ultrasound gel (Parker Aquasonic Clear) was applied to the probe before imaging. In the non-vitiligo cohort, we scanned the lower leg (calf muscle), the forearm (transverse and longitudinal radial artery), the upper arm (bicep muscle) and the neck (transverse carotid artery). In the vitiligo cohort, we scanned the same positions, with additional scans taken of specific vitiligo-affected areas where a contralateral non-affected area could be identified. Each position measured first with the colourimeter and then scanned for 30 seconds at a time and marked with a small sticker when necessary to identify the scan location for subsequent measurements. Three successive 10 s scans were acquired, from which values were aggregated (mean) for subsequent analyses. The whole imaging procedure was repeated three times to assess the reproducibility of the resulting images. To minimise the effects of patient and probe motion, three frames with minimal motion were chosen from each 30 second scan for subsequent analysis.

### Image reconstruction and processing

Photoacoustic images were reconstructed and analysed using the PATATO toolkit and custom Python code (*33*). Images were reconstructed using an iterative model-based image reconstruction algorithm (field of view 4 cm, 400 × 400 pixels). The time-series data was band-pass filtered before model-based reconstruction (5 kHz to 7 MHz). The filtered signals were divided by laser pulse energy to correct for per-pulse energy fluctuations. A qualitative depth-exponential weighting was applied to all images to improve the image contrast at depth for visualisation, however, this weighting was not used in the quantitative analysis.

Spectral unmixing was applied to the reconstructed data using the PATATO toolkit. The images were unmixed pixel-wise using literature values of the absorption spectra. Each pixel was expressed as a linear combination of deoxyhaemoglobin (Hb) and oxyhaemoglobin (HbO_2_) using NumPy’s matrix pseudo-inverse function. Total haemoglobin (THb = HbO_2_ + Hb) and blood oxygenation estimates (sO_2_^EST^ = HbO_2_ / THb) were obtained from the unmixed coefficients.

Reconstructed ultrasound images were provided by the imaging system and were acquired in sequence with the photoacoustic images. Ultrasound images from each scan were used to identify a single frame in each 10 s scan that had minimal motion between the first and last wavelength of the PAI frame. This frame was used in the subsequent analyses. Polygonal regions of interest were drawn around the skin surface (epidermis), arteries, muscles and subcutaneous fat regions using ultrasound images as a reference. These were converted into binary masks and applied to the photoacoustic images. Unless specified, thresholds were applied to sO_2_^EST^ quantification when masks were applied; pixels with sO_2_^EST^ greater than 1 or smaller than 0 were ignored. Mean values of each metric (unmixed or single wavelength, as specified in the results) were calculated across the region of interest. Negative pixel values were excluded when calculating the mean spectra.

### Optical modelling and fluence correction

We simulated light fluence in a tissue model using a Monte-Carlo simulation in Monte Carol eXtreme (MCX) (*34*). The model was defined on a 1000 × 1000 × 1000 grid with pixel size 60 µm. Scattering properties were defined using typical values seen in soft tissue throughout, 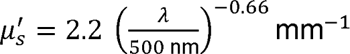, g = 0.9 (*3*). Three layers were used in the tissue model. On top, a 60 µm epidermis was modelled, with melanosome volume fraction varying logarithmically from 0.02 to 0.4 in 6 steps. Below, a 1 mm-thick dermis layer was modelled, with 0.2 % blood volume fraction and 70 % oxygenation. The rest of the simulation volume comprised a background layer with 70 % blood oxygenation and 2.5 % blood volume fraction (*11*). Finally, a blood vessel was added with radius 1.5 mm, 100 % blood oxygenation and 100 % blood volume fraction, at a depth of 5 mm.

The Monte-Carlo simulation was run with 10^7^ photons per simulation, at 41 wavelengths from 700 nm to 900 nm inclusive. We used an illumination geometry which approximates the clinical PAI system used in this study. The system consists of a single optical fiber with a beam divergence of 8.66°, 43.2 mm above the surface of the probe, directed at an angle of 22.4° from the imaging plane, intersecting the imaging plane 2.8 mm outside of the probe. All simulations were run on a computer with 2 NVIDIA Quadro 8000 GPUs (48 GB RAM each), Intel Xeon Gold 6230 CPU at 2.10 GHz (40 cores), and 256 GB RAM.

The fluence across the blood vessel region was averaged and saved for each wavelength and melanosome volume fraction. A lookup table for correction was then constructed by normalising all the factors by the wavelength-dependent fluence for the lowest melanosome volume fraction (0.02). The individual typology angle (ITA) measurements from colourimeter were related to melanosome volume fraction of the simulation by a previously-published adding-doubling diffuse reflectance simulation (*11*). The previous study estimated ITA as a function of melanosome volume fraction in the same simulation conditions as used here for the Monte-Carlo model. A quadratic curve was fit to the points from the previous study and used to estimate the appropriate melanosome volume fraction from the experimental ITA value.

To correct experimental data, we interpolated the normalised fluence correction factor using a cubic spline interpolation across wavelength and melanosome volume fraction using the SciPy class RegularGridInterpolator in Python. Each entire experimental photoacoustic image was then divided by the factor for the corresponding wavelength, after which the standard analysis and linear unmixing approach was taken.

### Melanin absorption plane-wave ultrasound reconstruction

Melanin ultrasound (mUS) images were generated from the raw photoacoustic time series data by adapting a pulse-echo plane-wave ultrasound reconstruction algorithm (*30*). At shorter optical wavelengths in subjects with darker skin, our photoacoustic time series acquisition resembles a plane-wave ultrasound imaging system like that described by Pham et al. (*30*), due to the strong, highly localised, and approximately planar optical absorption by melanin in the epidermis. Previous implementations of this algorithm used a planar ultrasound detection in the same position as the plane wave was generated. In this study, however, an arc-shaped ultrasound transducer array was used which was spatially offset from the skin surface, the source of the plane waves. The plane-wave reconstruction algorithm described by Cheng et al. (*35*) was therefore adapted to account for these two differences.

### Theoretical background

In two dimensions, the conventional plane-wave reconstruction algorithm aims to recover a B-mode-like ultrasound image, *a(x,y)*, which resembles the spatial distribution of ultrasound scatterers under a Born approximation. The method assumes that the acoustic field can be expressed as linear sum of plane waves, and that the dispersion relation 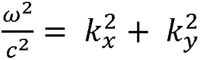 applies (*35*). Given the dispersion relation, any acoustic field can therefore be expressed as a function of any two of *k_x_, k_y_*, and *ω*. For an incident plane-wave 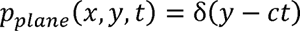 and resultant acoustic pressure time series measured by some detectors at 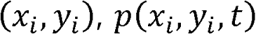, the algorithm proceeds by computing the Fourier transform of the pressure field, *P*(*k_x_,k_y_*) and applying the mapping 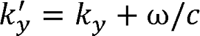. The mapping gives the Fourier transform of the scattering distribution, 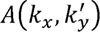. By applying the inverse Fourier transform, the scattering distribution can be recovered, *a*(*x,y*).

With planar ultrasound detection (detectors along the line *y* = 0), the algorithm proceeds by applying a two-dimensional Fourier transform to the measured time series *p*(*x*,*y* = 0,*t*) along the *x* and *t* axes, giving *P*(*k_x_,ω*). Through the dispersion relation, this can be mapped to *P*(*k_x_,k_y_*). Applying the second mapping, 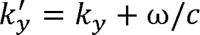, to get 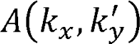, and inverse Fourier transforming, the scattering distribution, *a*(*x,y*) can be recovered. To account for a spatial offset between the skin surface (source of incident plane waves) and the ultrasound transducers, *P*(*k_x_,k_y_*) is multiplied by a phase factor exp(*ik_y_δ_y_*) before mapping to 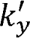 coordinates and 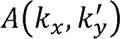 is multiplied by the phase factor 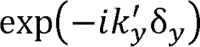 prior to the final inverse Fourier transform, where *δ_y_* is the perpendicular distance between the skin surface and the transducer array.

For the circular arc-shaped system used in this study, a plane wave ultrasound reconstruction algorithm was developed by analogy with the linear array. The Fourier transform of the pressure field, *P*(*k_x_,k_y_*) was computed, as described elsewhere (*36*). The algorithm then proceeds as above: multiply *P*(*k_x_,k_y_*), apply the mapping exp(*ik_y_δ_y_*) multiply by 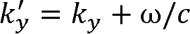 to get 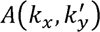, multiply by 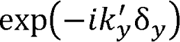, and apply and inverse Fourier transform to give the ultrasound scattering distribution *a*(*x,y*).

### Implementation

The melanin absorption ultrasound reconstruction algorithm was implemented using custom-written code in Python and is publicly available https://github.com/BohndiekLab/PAISKINTONE. As with photoacoustic image reconstruction, a Butterworth bandpass filter was applied to the raw photoacoustic time series data between 5 kHz and 7 MHz using the SciPy functions butter (order 5) and filtfilt (even padding with length 100).

To isolate the scattered ultrasound waves, the non-scattered wave generated by the skin surface was subtracted from the filtered raw time series data. To calculate the non-scattered wave, the forward photoacoustic model was applied to the photoacoustic image with pixels outside the skin surface set to zero. The subtracted and filtered timeseries data was then used as input for the plane wave reconstruction algorithm described above.

To compute the Cartesian Fourier transform, *P*(*k_x_,k_y_*) from the (filtered, subtracted) arc-shaped plane-wave timeseries data, *P*(θ*,t*), a Fourier-based method was used. First, a smooth crop of the time-series data was made. Then, the time-series was interpolated onto a grid covering all 2π radians, with zeros used in place of missing detector angles. A two-dimensional Fourier transform was applied along the angle and time dimensions. A Hankel function weighting was applied, the data Fourier transformed again along the angle dimension, before interpolation onto a Cartesian grid to give *P*(*k_x_,k_y_*). The plane-wave ultrasound reconstruction algorithm then proceeded as described above, using the Python packages NumPy and SciPy.

### Validation using acoustic simulations

To validate the adapted algorithm described above, two dimensional k-space plane-wave ultrasound simulations were run in j-Wave, modelling the key components of ultrasound scattering reconstruction based on melanin absorption (*37*). A simulation grid was defined with a width and height of 256 pixels, the pixel width was 50 µm. The background speed of sound was 1540 m s^-1^, and the background density was 1000 kg m^-3^. The initial pressure distribution was defined to mimic skin surface absorption, with a single pixel-width horizontal structure of constant value. A sensor array with 256-point detectors was defined parallel to the skin surface, with a defined spatial offset δ_y_. The initial pressure in a simulated blood vessel region was defined, either with the same amplitude (low melanin limit) or 100 times less amplitude (high melanin limit). Simulations were run in two dimensions, with a 512 × 512 grid with 200 µm spacing. To simulate ultrasound scattering structures, inhomogeneities in the tissue density and tissue speed of sound were introduced. Nine circular scattering structures were defined with radius 1.5 mm, 5 mm below the skin surface, evenly distributed on a square of side length 22.5 mm (Figure S8). Outside of the structures, the background density and speed of sound were multiplied by random noise sampled from a normal distribution with mean 1 and standard deviation 0.008. Inside the scattering circle structures, the density was sampled from a normal distribution with mean 1043 kg m^-3^ and standard deviation 61 kg m^-3^, and the speed of sound was sampled from a normal distribution with mean 1565 m s^-1^ and standard deviation 75 m s^-1^.

### Statistical analysis

Statistical analysis was carried out using the statsmodels library in Python. The relationship between continuous variables was assessed using a linear model in statsmodels, unless otherwise stated. Where a statistical test was applied to determine a dependence on skin pigmentation, the individual typology angle (ITA) was used as the dependent variable, and quoted p-values are the 2-tailed test. Where a linear model was not appropriate, a Spearman’s rank correlation test was applied using the spearmanr function in SciPy. Where stated, a t-test was applied using the SciPy function ttest_1samp.

The theoretical model of sO_2_^EST^ dependence on ITA was compared to an alternative model (sO_2_^EST^ = constant) by calculating the likelihood ratio, using the Akaike information criterion (AIC) for the two models. The log likelihood was calculated using the residual Values, *r_i_* as follows

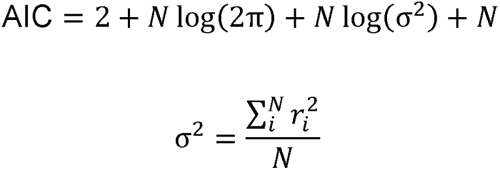

The likelihood ratio was then computed as follows:

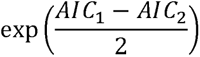

Where linear trend lines were not appropriate, locally weighted scatterplot smoothing (LOWESS) curves are included to guide the eye and highlight non-linear trends. LOWESS fits a smooth local polynomial regression line, allowing model fitting without the specification of a function. LOWESS was computed using the statsmodels lowess function. 75 % of the data was used when calculating each y-value (frac = 0.75 in lowess function), and 3 residual-based re-weightings were performed (it = 3 in lowess function). 95 % confidence intervals were calculated by bootstrapping, with 500 repeated re-samplings with replacement.

## Data Availability

Data associated with this manuscript will be made available on the University of Cambridge data repository upon publication in a peer-reviewed journal. Before that time, data produced are available upon reasonable request to the authors.

## Acknowledgments

This work was funded by: Cancer Research UK (SEB, TRE: C9545/A29580, AR: C64667/A27958). TRE and SEB also acknowledge funding from the Engineering and Physical Sciences Research Council (EP/X037770/1). This work was supported by the International Alliance for Cancer Early Detection, a partnership between Cancer Research UK [EDDAMC-2023/100003], Canary Center at Stanford University, the University of Cambridge, OHSU Knight Cancer Institute, University College London and the University of Manchester. This research was supported by NIHR Cambridge Biomedical Research Centre (NIHR203312). The views expressed are those of the authors and not necessarily those of the NIHR or the Department of Health and Social Care. JG acknowledges funding from the Walter Benjamin Stipendium of the Deutsche Forschungsgemeinschaft.

## Funding sources

Cancer Research UK C9545/A29580

Cancer Research UK C64667/A27958

Engineering and Physical Sciences Research Council UK EP/X037770/1

National Institute of Health Research UK NIHR203312

Cancer Research UK EDDAMC-2023/100003

Deutsche Forschungsgemeinschaft Walter Benjamin Stipendium

## Author contributions

Conceptualization: TRE, SEB, AR

Data curation: TRE, CL, AG

Formal analysis: TRE

Funding acquisition: SEB

Investigation: TRE, CL, AG

Methodology: TRE, BTC

Project administration: TRE, CL, AG, IM, SEB, AR

Software: TRE, JG

Supervision: SEB, AR, IM

Visualization: TRE

Writing – original draft: TRE, SEB, AR

Writing – review & editing: TRE, JG, BTC, SEB, AR

## Data and materials availability

https://github.com/BohndiekLab/PAISKINTONE. Data associated with this manuscript will be made available on the University of Cambridge data repository upon publication (DOI TBC).

