## Supplementary Information for "The confounding effects of skin colour in photoacoustic imaging"

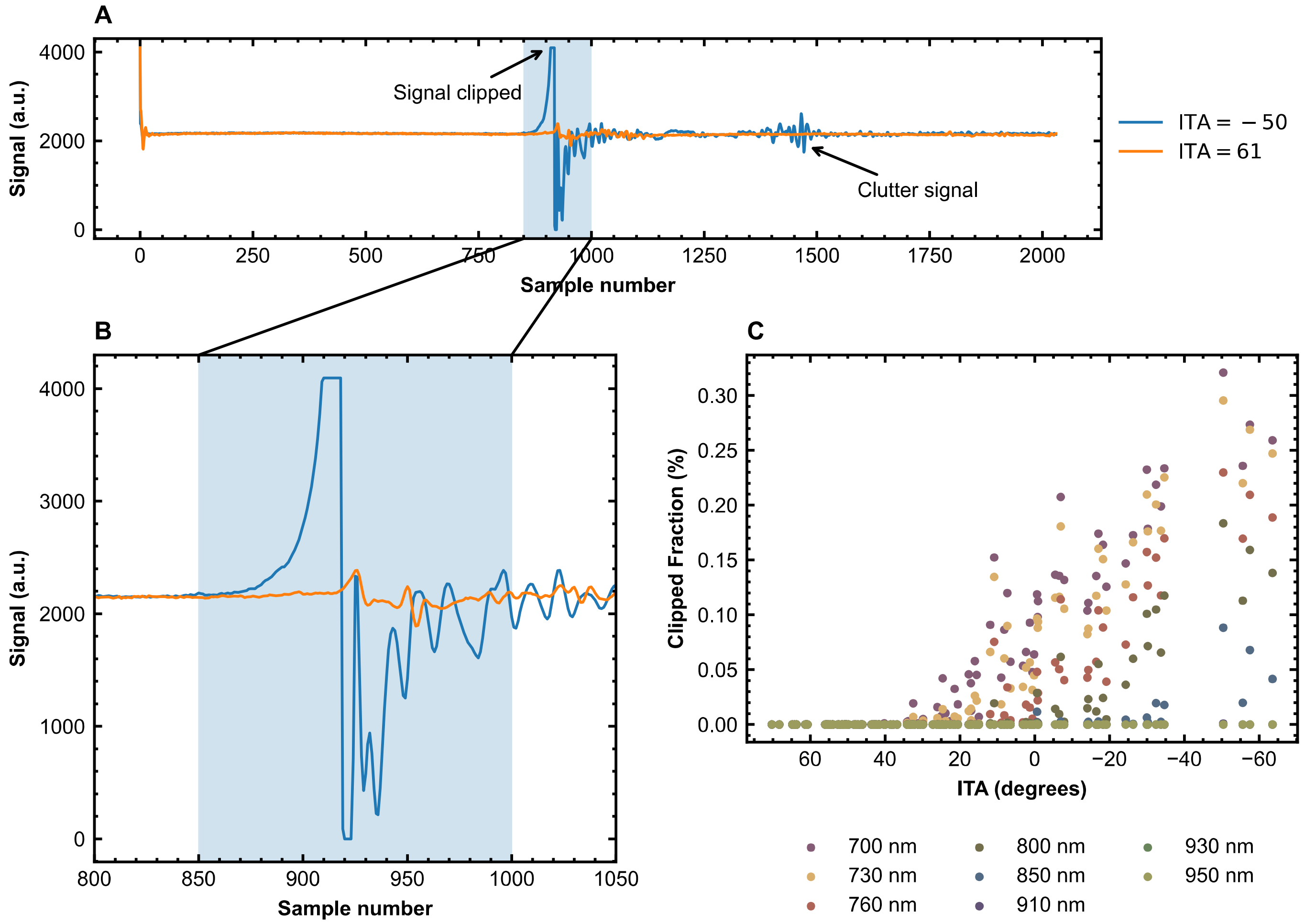


Figure S1: **Raw photoacoustic time-series data shows clipping in signal digitisation.** (**A** and **B**) Comparison of the raw time series data from a Fitzpatrick V subject, ITA = -50 (blue) and a Fitzpatrick II subject, ITA = 61 (orange). The signal can be seen to reach the limit of digitisation during the arrival of the skin signal. (**C**) The fraction of time samples that were clipped is shown as a function of wavelength and skin type.

| Study ID | Age range | Sex | Fitzpatrick type or vitiligo | BMI ($kg\,m^{-2}$) | ITA (°) |
|---|---|---|---|---|---|
| SKIN01 | 20-29 | Male | 2 | 25.5 | 54.1 |
| SKIN02 | 30-39 | Male | 2 | 29.9 | 38.5 |
| SKIN03 | 40-49 | Male | 3 | 27.5 | 17.0 |
| SKIN04 | 20-29 | Female | 3 | 22.0 | 31.1 |
| SKIN05 | 20-29 | Female | 3 | 21.5 | 36.8 |
| SKIN06 | 70-79 | Female | 2 | 21.6 | 27.2 |
| SKIN07 | 30-39 | Male | 3 | 27.7 | 33.8 |
| SKIN08 | 30-39 | Male | 4 | 24.6 | −19.1 |
| SKIN09 | 50-59 | Female | 4 | 21.9 | 8.1 |
| SKIN10 | 60-69 | Female | 2 | 26.6 | 37.8 |
| SKIN11 | 50-59 | Female | 2 | 18.5 | 21.6 |
| SKIN12 | 70-79 | Female | 1 | 19.1 | 41.8 |
| SKIN13 | 20-29 | Male | 2 | 25.2 | 39.7 |
| SKIN14 | 30-39 | Female | 4 | 27.2 | 0.6 |
| SKIN15 | 30-39 | Female | 4 | 24.2 | 22.6 |
| SKIN16 | 70-79 | Female | 1 | 20.2 | 40.2 |
| SKIN17 | 20-29 | Female | 3 | 19.7 | 30.7 |
| SKIN18 | 30-39 | Female | 3 | 23.2 | 26.1 |
| SKIN19 | 30-39 | Female | 5 | 27.5 | −63.5 |
| SKIN20 | 50-59 | Male | 1 | 23.1 | 42.5 |
| SKIN21 | 20-29 | Female | 1 | 22.0 | 47.2 |
| SKIN22 | 20-29 | Male | 4 | 22.6 | −5.5 |
| SKIN23 | 20-29 | Female | 1 | 21.9 | 50.9 |
| SKIN24 | 50-59 | Female | 1 | 20.8 | 46.1 |
| SKIN25 | 20-29 | Male | 4 | 19.6 | 6.5 |
| SKIN26 | 60-69 | Male | 5 | 24.5 | −6.6 |
| SKIN28 | 70-79 | Female | Vitiligo | 28.7 | 44.9 |
| SKIN29 | 40-49 | Male | 6 | 22.5 | −33.8 |
| SKIN30 | 40-49 | Female | Vitiligo | 20.3 | 51.2 |
| SKIN31 | 30-39 | Male | 6 | 20.9 | −55.6 |
| SKIN32 | 40-49 | Male | 5 | 25.1 | −0.8 |
| SKIN33 | 40-49 | Male | Vitiligo | 28.5 | 47.6 |
| SKIN34 | 60-69 | Female | Vitiligo | 28.3 | 39.2 |
| SKIN35 | 40-49 | Female | 6 | 26.4 | −39.3 |
| SKIN36 | 70-79 | Male | Vitiligo | 20.2 | 14.0 |
| SKIN37 | 60-69 | Female | Vitiligo | 23.9 | 37.5 |
| SKIN38 | 20-29 | Female | 6 | 27.5 | −67.7 |
| SKIN39 | 20-23 | Male | 5 | 24.5 | −15.2 |
| SKIN40 | 20-29 | Male | 6 | 25.8 | −60.4 |
| SKIN41 | 20-29 | Male | 6 | 19.6 | −61.1 |
| SKIN42 | 20-29 | Female | 5 | 20.7 | −16.9 |
| SKIN43 | 30-39 | Female | 5 | 20.2 | −25.6 |

Table S1: **Summary of all participants recruited to the study.** Note: SKIN27 was consented but did not participate in the study because of equipment failure.

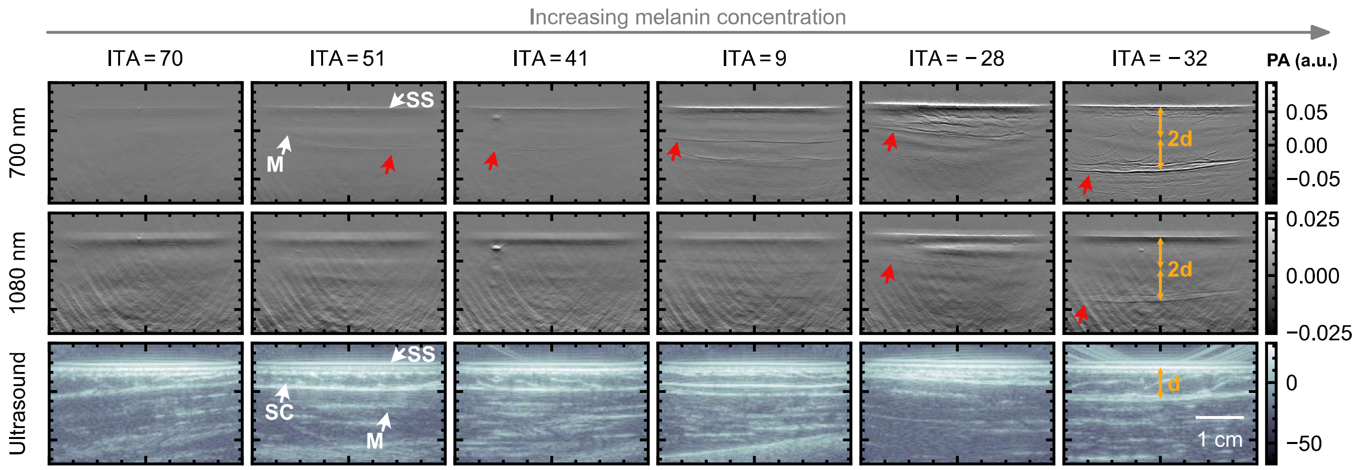


Figure S2: **Example photoacoustic and ultrasound images of the bicep muscle at representative skin tones and several wavelengths.** Key anatomical features are highlighted by a white arrow and acoustic reflection artefacts are highlighted by a red arrow (M = muscle, SS = skin surface, SC = subcutaneous fat). Orange arrows show that the artefact depth is twice the depth from the skin surface to muscle-fat interface. A qualitative exponential signal intensity depth compensation is applied to enhance the visibility of deeper structures.

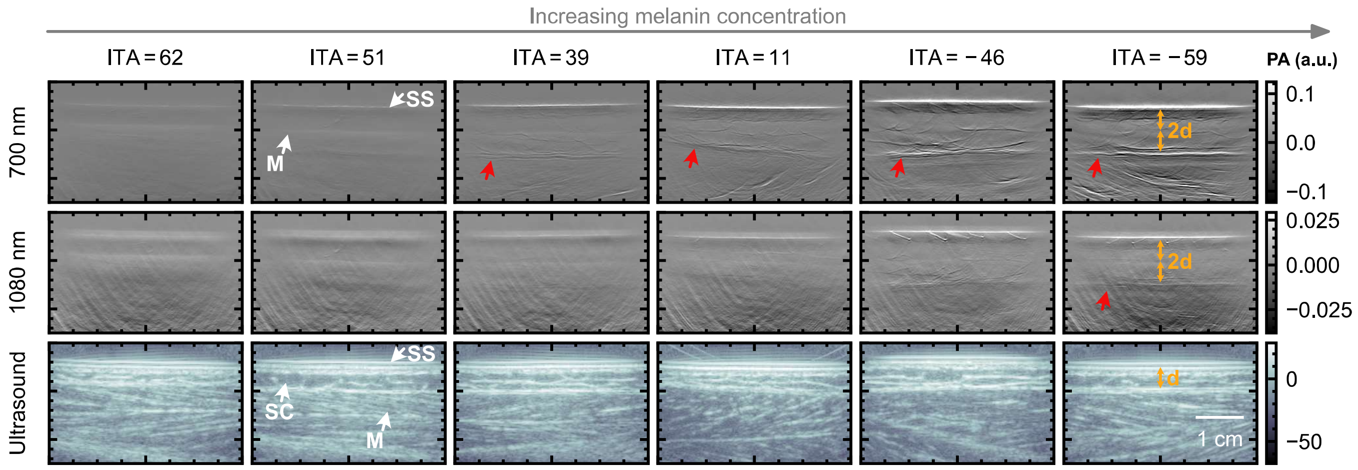


Figure S3: **Example photoacoustic and ultrasound images of the calf muscle at representative skin tones and several wavelengths.** Key anatomical features are highlighted by a white arrow and acoustic reflection artefacts are highlighted by a red arrow (M = muscle, SS = skin surface, SC = subcutaneous fat). Orange arrows show that the artefact depth is twice the depth from the skin surface to muscle-fat interface. A qualitative exponential signal intensity depth compensation is applied to enhance the visibility of deeper structures.

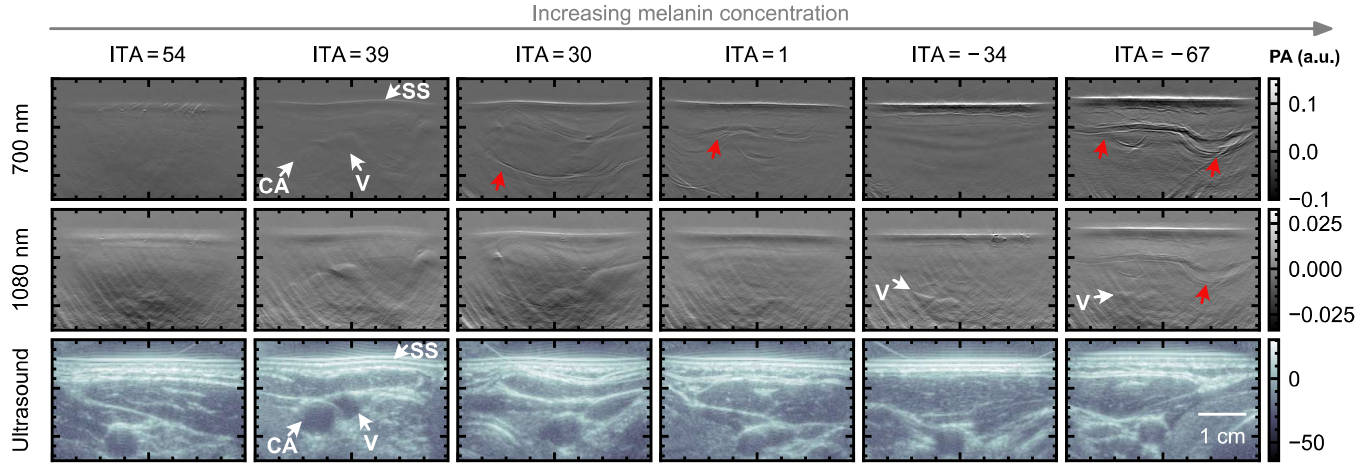


Figure S4: **Example photoacoustic and ultrasound images of the carotid artery at representative skin tones and several wavelengths.** Key anatomical features are highlighted by a white arrow and acoustic reflection artefacts are highlighted by a red arrow (V = vein, CA = carotid artery, SS = skin surface). A qualitative exponential signal intensity depth compensation is applied to enhance the visibility of deeper structures.

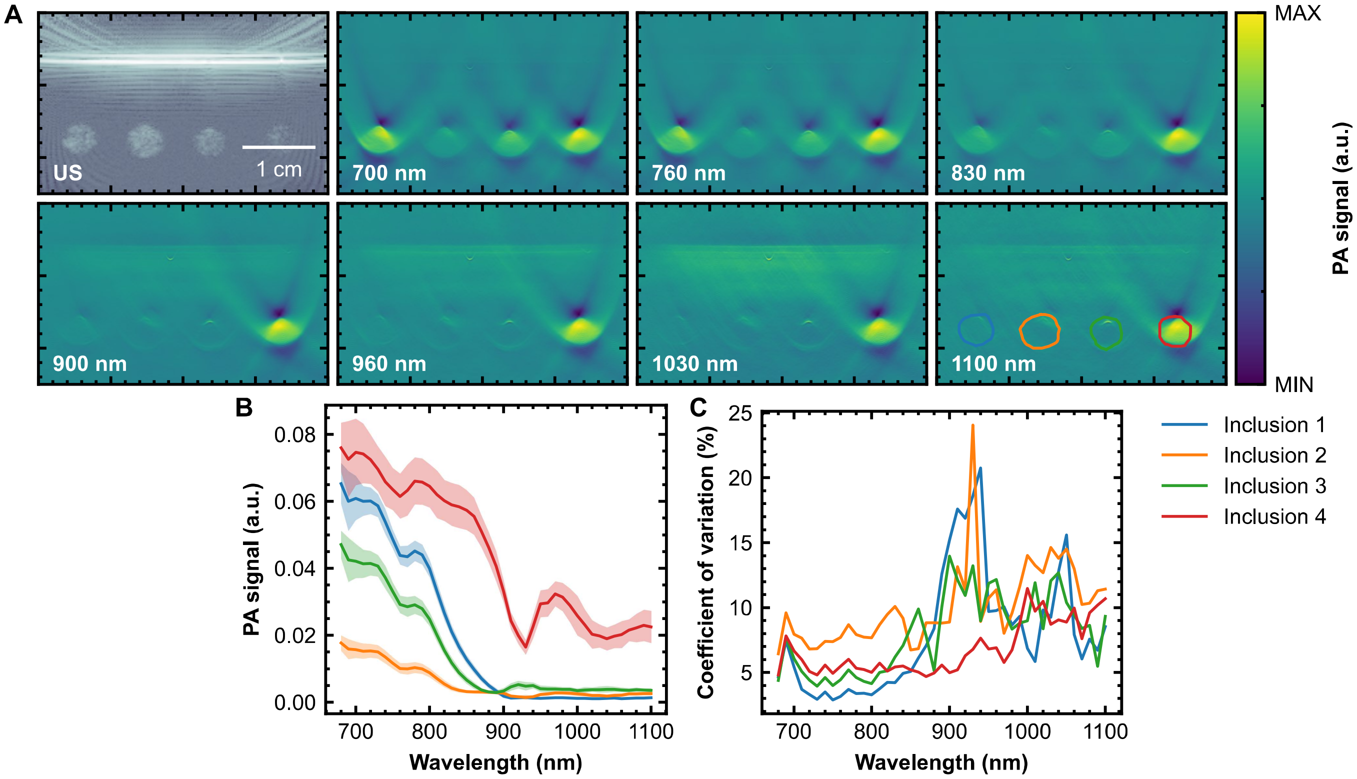


Figure S5: **Characterisation of phantom imaging reproducibility over the course of the study.** (**A**) Example ultrasound image of the test phantom. (**B**) Mean photoacoustic spectrum across (solid line) and standard deviation (shaded area) in each phantom inclusion, across phantom scans. (**C**) Coefficient of variation across all wavelengths for each phantom inclusion.

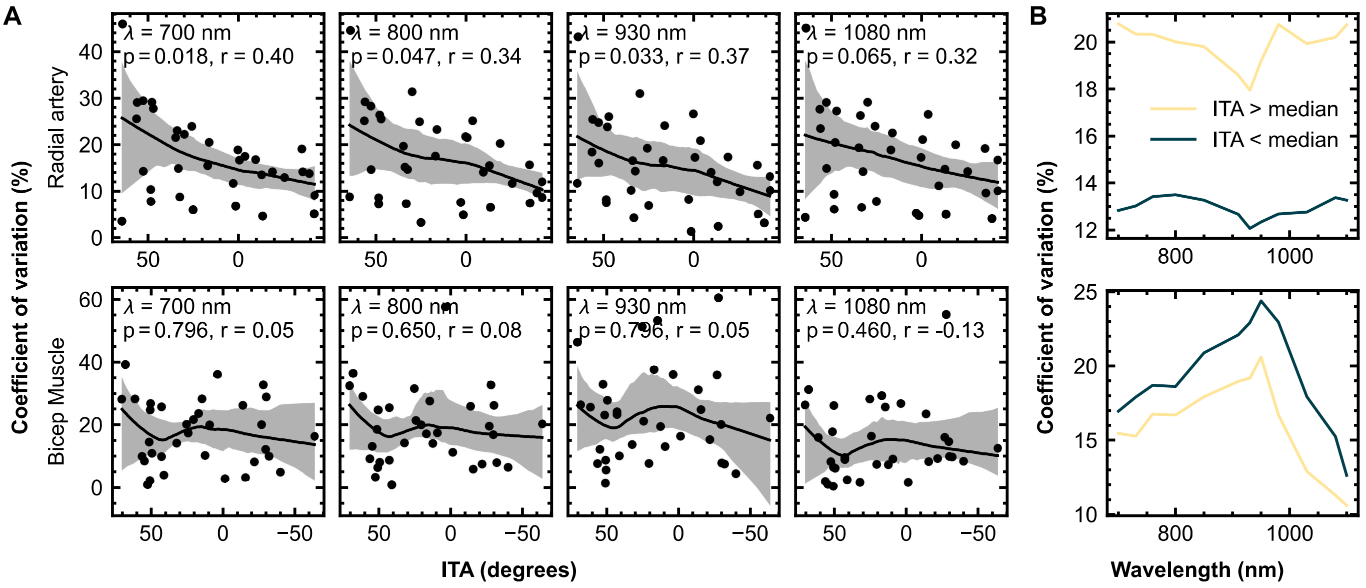


Figure S6: **Coefficient of variation in single wavelength measurements averaged across the radial artery and bicep muscle.** (**A**) A plot of coefficient of variation as a function of individual typology angle (ITA) at several wavelengths for each region of interest, with a LOWESS smoothing curve and 95% confidence intervals shown as the shaded region, calculated by bootstrapping. Spearman's rank correlation coefficient r and p values are shown. (**B**) The mean coefficient of variation across all wavelengths is plotted for the half cohort of participants with lighter skin (ITA$>$ median) and darker skin (ITA $<$ median) in the radial artery and bicep muscle.

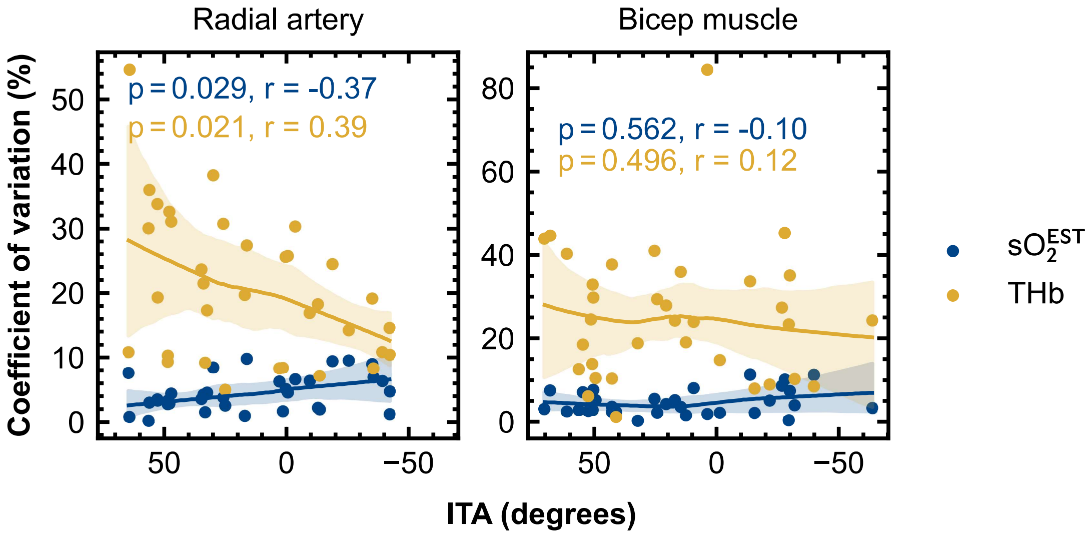


Figure S7: **Coefficient of variation in linear unmixing quantities, $\mathrm{sO}_2^{\mathrm{EST}}$ and THb, averaged across the radial artery and bicep muscle.** A plot of coefficient of variation as a function of individual typology angle (ITA) for each region of interest. Spearman's rank correlation coefficient r and p values are shown. LOWESS smoothing is shown with 95% confidence intervals shaded.

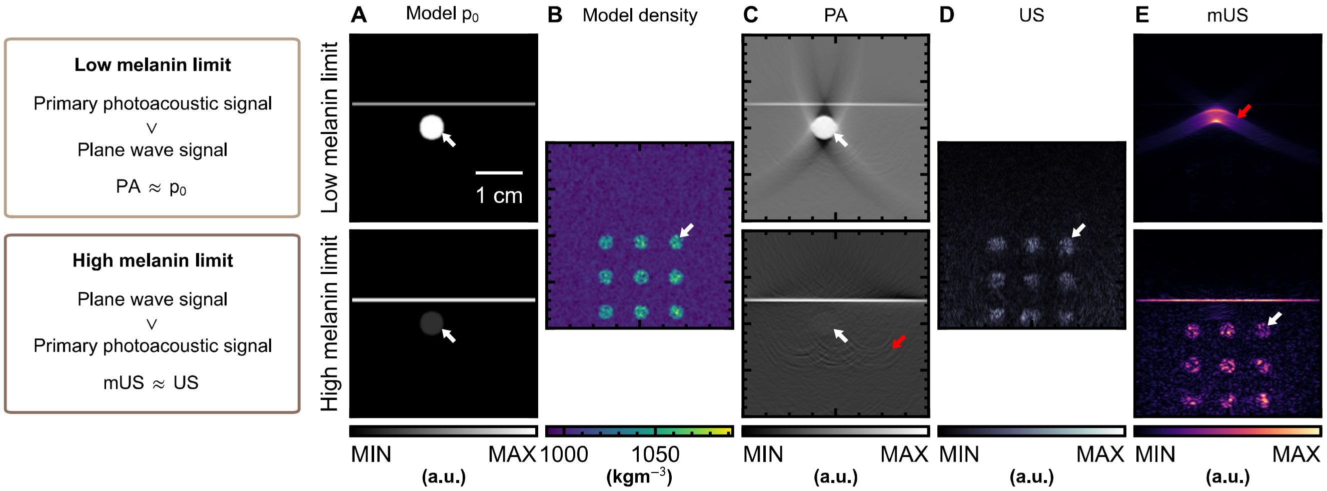


Figure S8: **Acoustic simulations in the low and high melanin concentration limits validate the melanin absorption ultrasound (mUS) reconstruction method.** Initial pressure (**A**) and tissue density (**B**) parameters used in the acoustic simulations; bright circular structures represent a blood vessel ($p_0$) and generic ultrasound scatterers (density) respectively. (**C**) Reconstructed simulated photoacoustic (PA) images show the skin surface, the blood vessel (white arrow) and artefacts (red arrows). (**D**) Reconstructed simulated ultrasound (US) images. (**E**) Reconstructed melanin ultrasound images based on plane wave reconstruction. Artefacts in each image are highlighted with a red arrow. The blood vessel is indicated with a white arrow in the PA image and ultrasound scatterers are highlighted with a white arrow in US and mUS images. *Note: the colour scales have been manually adjusted to highlight key artefacts and image features.*

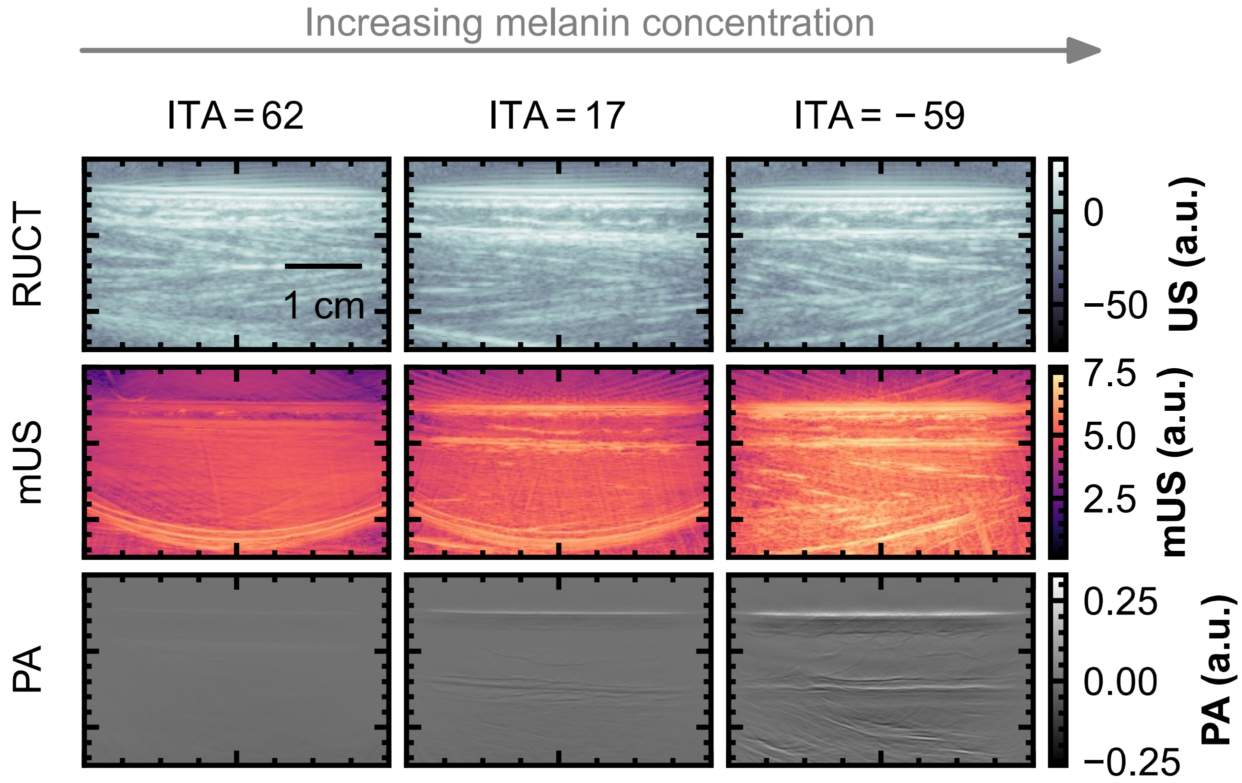


Figure S9: **Melanin absorption ultrasound (mUS) images in the calf muscle.** Conventional, reflection ultrasound computed tomography (RUCT) images, compared to melanin ultrasound (mUS) images from plane wave ultrasound reconstruction, and photoacoustic (PA) images in scans of the calf muscle in several different subjects with a range of skin tones.

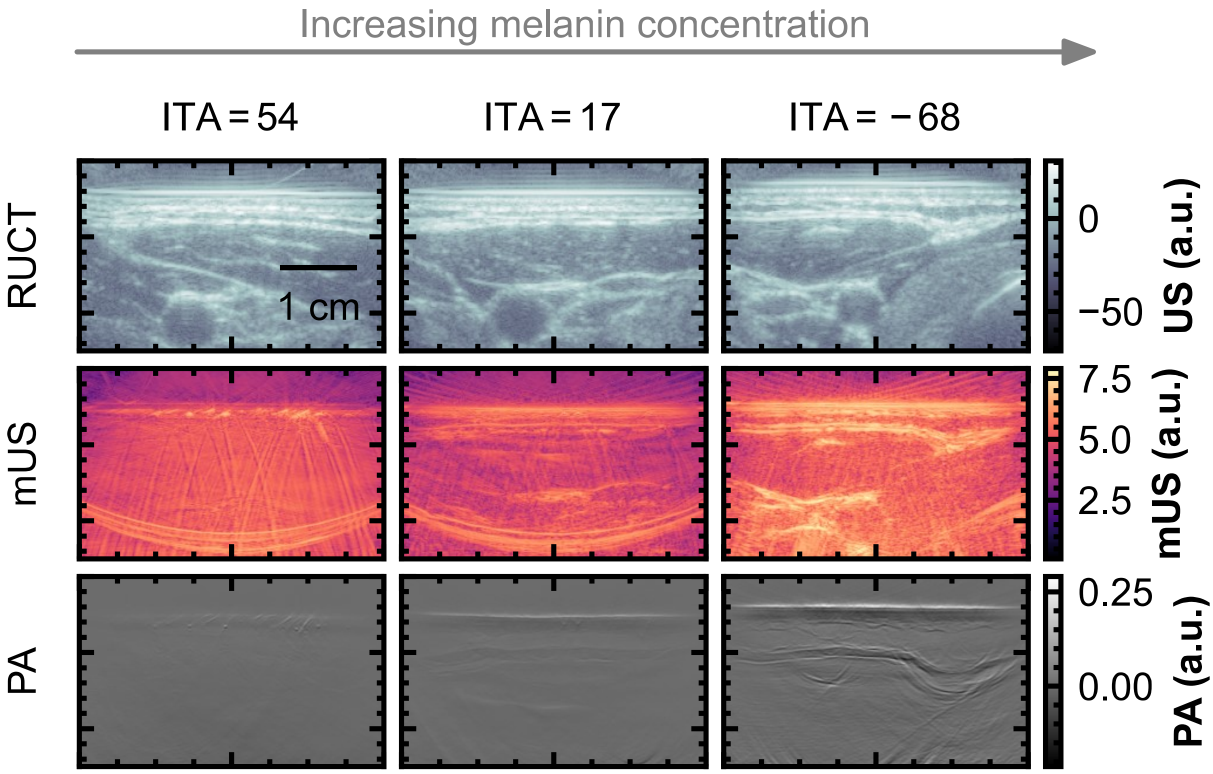


Figure S10: **Melanin absorption ultrasound (mUS) images of the carotid artery.** Conventional, reflection ultrasound computed tomography (RUCT) images, compared to melanin ultrasound (mUS) images from plane wave ultrasound reconstruction, and photoacoustic (PA) images in scans of the carotid artery in the neck in several different subjects with a range of skin tones.
